# Meta-Analysis of Rare Cancers Leveraging Clinically Ascertained Cohorts Reveals Novel Germline Susceptibility Loci

**DOI:** 10.64898/2026.04.16.26350975

**Authors:** Shaye Carver, Tomin Perea-Chamblee, Kodi Taraszka, Intae Moon, Xinran Yu, Yi Ding, Jian Carrot-Zhang, Alexander Gusev

## Abstract

Genome-wide association studies (GWAS) have advanced the understanding of germline susceptibility in common cancers, yet rare malignancies remain underexplored due to limited sample sizes. To address this gap, we conducted large-scale GWAS across 20 rare cancer types and meta-analyzed results from three cohorts: two clinically sequenced cancer center cohorts and an independent population biobank, comprising over 480,000 individuals. We identified nine novel genome-wide significant susceptibility loci with moderate to large effect sizes that replicated across cohorts in eight rare malignancies, including myelodysplastic syndromes (MDS), germ cell tumors, gastrointestinal stromal tumor (GIST), gastrointestinal neuroendocrine tumors, anal cancer (ANSC), non-melanoma skin cancer, mesothelioma, and hepatobiliary cancer. Among the strongest associations were loci in MDS near *API5* (OR = 2.21, *p* = 1.06×10^−8^), in GIST near *SLC6A18* and *TERT* (OR = 1.91, *p* = 8.20×10^−50^), and in ANSC near HLA-DQA2 (OR = 1.58, *p* = 5.50×10^−18^). The GIST risk variant was enriched in tumors harboring somatic KIT mutations (OR = 2.21, p = 6.5×10^−4^) and was associated with worse survival among carriers with KIT-mutant tumors (hazard ratio = 4.06, p = 0.015), implicating germline–somatic interplay in tumor initiation and progression. The ANSC risk variant was associated with HPV infection (OR = 1.44, p = 3.19×10^−5^), supporting a host–viral interaction in HPV-driven tumorigenesis. The MDS risk variant at the *API5* locus was associated with altered neutrophil counts, suggesting a role in hematopoietic dysregulation in disease pathogenesis. We further identified novel, independent associations with mesothelioma, GIST, and hepatobiliary cancer at the 5p15.33 locus encompassing *TERT*, consistent with pleiotropic genetic effects at a core telomere-maintenance gene. Collectively, these findings demonstrate that integrating clinically ascertained sequencing cohorts with population biobanks substantially enhances germline discovery in rare cancers, enabling identification of high-confidence susceptibility loci and facilitating downstream biological interpretation through linked somatic, viral, and clinical data. This framework provides a scalable approach for characterizing inherited susceptibility across diverse rare malignancies.

## Introduction

Genome-wide association studies (GWAS) have identified hundreds of germline variants associated with the risk of many common cancers^1–4^. However, for rare cancer types, the difficulty of recruiting sufficiently large patient cohorts has hindered the identification of germline risk variants, limiting our ability to characterize their genetic architecture and constraining the development of targeted treatment options informed by their underlying genetic basis^5–8^. Despite their rarity, these cancers collectively account for a large fraction of the overall cancer burden, with considerable associated morbidity and mortality^7,9,10^. For most rare malignancies, limited sample sizes have precluded estimation of even basic parameters such as heritability or polygenicity^11,12^. Consequently, it remains unknown whether rare cancers are largely heritable, follow Mendelian-like inheritance driven by high-penetrance variants, or exhibit a polygenic architecture driven by common variation.

Advances in sequencing technologies and analytical methods now enable GWAS in routinely collected clinical samples^13^. Clinical tumor sequencing is performed at scale as part of routine oncologic care, with hundreds of thousands of tumors having been sequenced to date across large health systems^14–17^. Although these efforts have primarily focused on identifying somatic driver alterations and somatic biomarkers for clinical decision support^14,17–21^, we have previously shown that common germline variants can be imputed directly from such targeted tumor sequencing data^13^ and leveraged for downstream genetic analyses including GWAS^22,23^. This framework enables the study of germline mechanisms in cancers that are otherwise underpowered or infeasible to investigate using traditional cohorts or biobank-based approaches. In addition, leveraging cancer sequencing cohorts provides access to richly annotated tumor, treatment, and outcome data, enabling mechanistic and clinical dissection of germline risk loci that is often not possible in population-scale biobanks. The resulting multi-modal data integration provides a unique opportunity to systematically characterize the germline genetic architecture of rare cancers in the context of tumor evolution, disease progression, and clinical aggressiveness.

In this study, we systematically investigated the germline genetic architecture of 20 rare cancers, motivated by prior evidence that jointly analyzing multiple rare cancers can increase power for discovery and uncover shared genetic architecture^1^. Leveraging approximately 40,000 cases across these malignancies and an average of over 360,000 controls per cancer type (**Figure 1A–B; Supplementary Table 1**), we integrated large-scale cancer sequencing cohorts with population-based biobank data to conduct a meta-analysis that identified nine novel genome-wide significant germline associations across eight cancers (**Supplementary Tables 2 and 3**). The availability of detailed clinical data further enabled analyses of germline–somatic interactions, downstream clinical outcomes, and environmental exposures. Together, these results demonstrate that rare cancers can be driven by large-effect common germline variants, reveal both shared susceptibility across rare cancers and distinct cancer-specific risk loci, and establish a generalizable framework for genetic discovery in rare malignancies.

**Figure 1:**
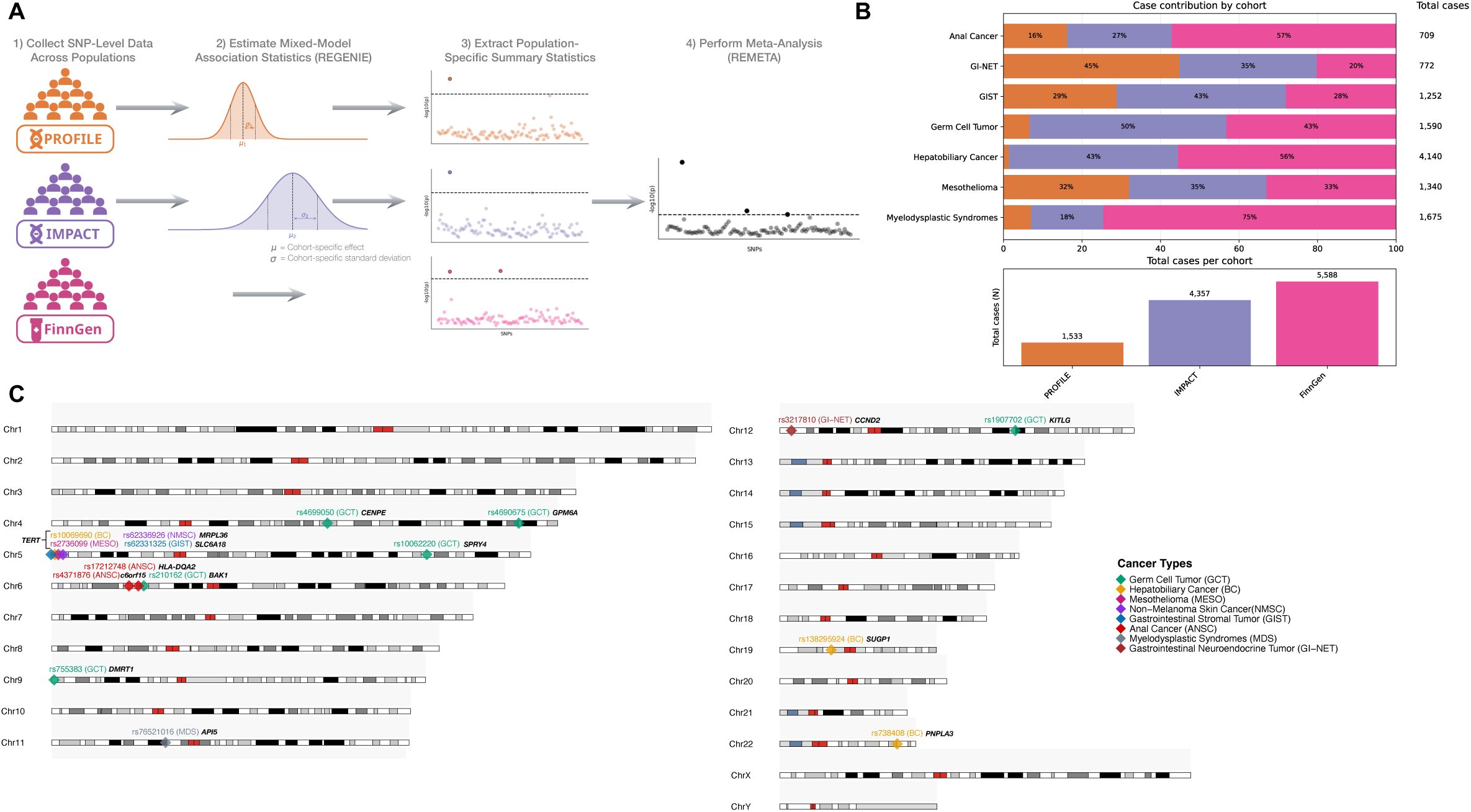
Study overview, cohort composition, and discovery of genome-wide significant loci. (**A**) Schematic of the meta-analysis framework integrating three cohorts: PROFILE and IMPACT (tumor sequencing cohorts) and FinnGen (a population biobank). Genome-wide association analyses were performed in PROFILE and IMPACT using REGENIE, combined with publicly available FinnGen summary statistics, and meta-analyzed using REMETA. (**B**) Overview of rare cancers in the meta-analysis with novel genome-wide significant loci. For each cancer type, the proportion of cases contributed by each cohort is shown alongside the total number of cases (top), and the total number of cases contributed by each cohort across all cancers is summarized (bottom). (**C**) Karyotype plot showing the genomic distribution of novel genome-wide significant loci identified across all cancers. Diamonds denote lead variants (rsIDs), colored by cancer type, with adjacent labels (black text) indicating the nearest gene.

## Results

### Study Cohorts and Meta-Analysis Framework

We evaluated germline genetic associations across rare cancers using three large cohorts: two clinically curated cancer sequencing cohorts from Dana-Farber Cancer Institute (PROFILE; n = 35,260) and Memorial Sloan Kettering Cancer Center (IMPACT; n = 68,249), and the population-based FinnGen biobank (n ≈ 380,000) (**Figure 1A**). In PROFILE and IMPACT, cancer diagnoses were confirmed by institutional pathology review, and individuals with other cancer types served as controls. In FinnGen, cancer status was defined using electronic health record and billing codes, and cancer-free individuals were used as controls.

We analyzed 20 malignancies classified as rare, defined as affecting fewer than 1% of individuals in PROFILE or exhibiting low population-level incidence according to American Cancer Society statistics (**Supplementary Table 1**).

For each cancer type, we implemented a staged analytical framework. GWAS were first conducted independently in PROFILE and IMPACT using REGENIE^24^, a whole-genome regression framework that approximates a mixed-effects model to account for population structure, relatedness, and technical covariates (see **Methods**). Associations replicating across the two clinical cohorts were then combined in a two-cohort meta-analysis using REMETA^25^, which performs summary statistic–based effect-size and p-value meta-analysis of single variants. Variants demonstrating additional evidence of replication in FinnGen were subsequently incorporated into a three-cohort meta-analysis within the same framework using publicly available summary statistics.

Across this framework, cancer types yielding novel genome-wide significant associations included myelodysplastic syndromes (1,675/103,083), germ cell tumor (1,590/259,277), gastrointestinal stromal tumor (GIST; 1,252/481,303), gastrointestinal neuroendocrine tumors (772/102,893), anal cancer (ANSC; 709/481,954), non-melanoma skin cancer (29,285/480,372), mesothelioma (1,340/481,360), and hepatobiliary cancer (4,140/480,372) (**Figure 1B; Supplementary Table 3**). The incorporation of clinically curated cancer cohorts substantially increased case counts relative to FinnGen biobank data alone, with increases of 1.8-fold for ANSC, 3.6-fold for GIST, 2.3-fold for germ cell tumors, 3.0-fold for mesothelioma, and 1.8-fold for hepatobiliary cancer (**Supplementary Table 3**).

### Novel Germline Risk Loci Identified by Cross-Cohort Meta-Analysis

Across 20 rare cancer types, our cross-cohort meta-analytic GWAS framework, spanning clinically curated cancer cohorts and a population-based biobank, both replicated established susceptibility loci and identified novel germline risk associations. In the three-cohort meta-analysis, we reproduced seven previously reported genome-wide significant loci across germ cell and hepatobiliary cancer (**Supplementary Table 3**), including SPRY4, KITLG, the BAK1 region, DMRT1, and CENPE for germ cell tumors, and PNPLA3 and SUGP1 for hepatobiliary cancer. Analyses were restricted to variants that were directly genotyped or well imputed across all cohorts, and effect directions were largely concordant across the clinical cohorts and FinnGen. The robust recovery of known associations supports the validity of our analytical framework.

In addition, we identified nine novel genome-wide significant germline associations across eight cancers (**Figure 1C**). We focused subsequent analyses on loci that replicated across both clinically curated cohorts (PROFILE and IMPACT) or across all three cohorts (PROFILE, IMPACT, and FinnGen), and that have not been previously reported in large-scale cancer GWAS. All identified variants were common in the general population (allele frequency > 0.05 in gnomAD), with one exception (rs76521016; AF = 0.043; **Supplementary Table 4**). These loci demonstrated moderate to large effect sizes and consistent directions of effect across cohorts, supporting their robustness. Below, we describe these findings in descending order of effect size, prioritizing estimates from the three-cohort meta-analysis where available and otherwise reporting results from the two-cohort clinical meta-analysis.

The largest effect was observed in myelodysplastic syndromes (1,675 cases), where rs76521016-C upstream of *API5* was significantly associated with risk in the PROFILE–IMPACT meta-analysis (OR = 2.21; *p* = 1.06×10^−8^), with consistent effects across both clinical cohorts (**Figure 2A; Supplementary Table 3**). *API5* encodes an inhibitor of apoptosis, suggesting a potential mechanistic link to altered survival of dysplastic hematopoietic cells^26^.

**Figure 2:**
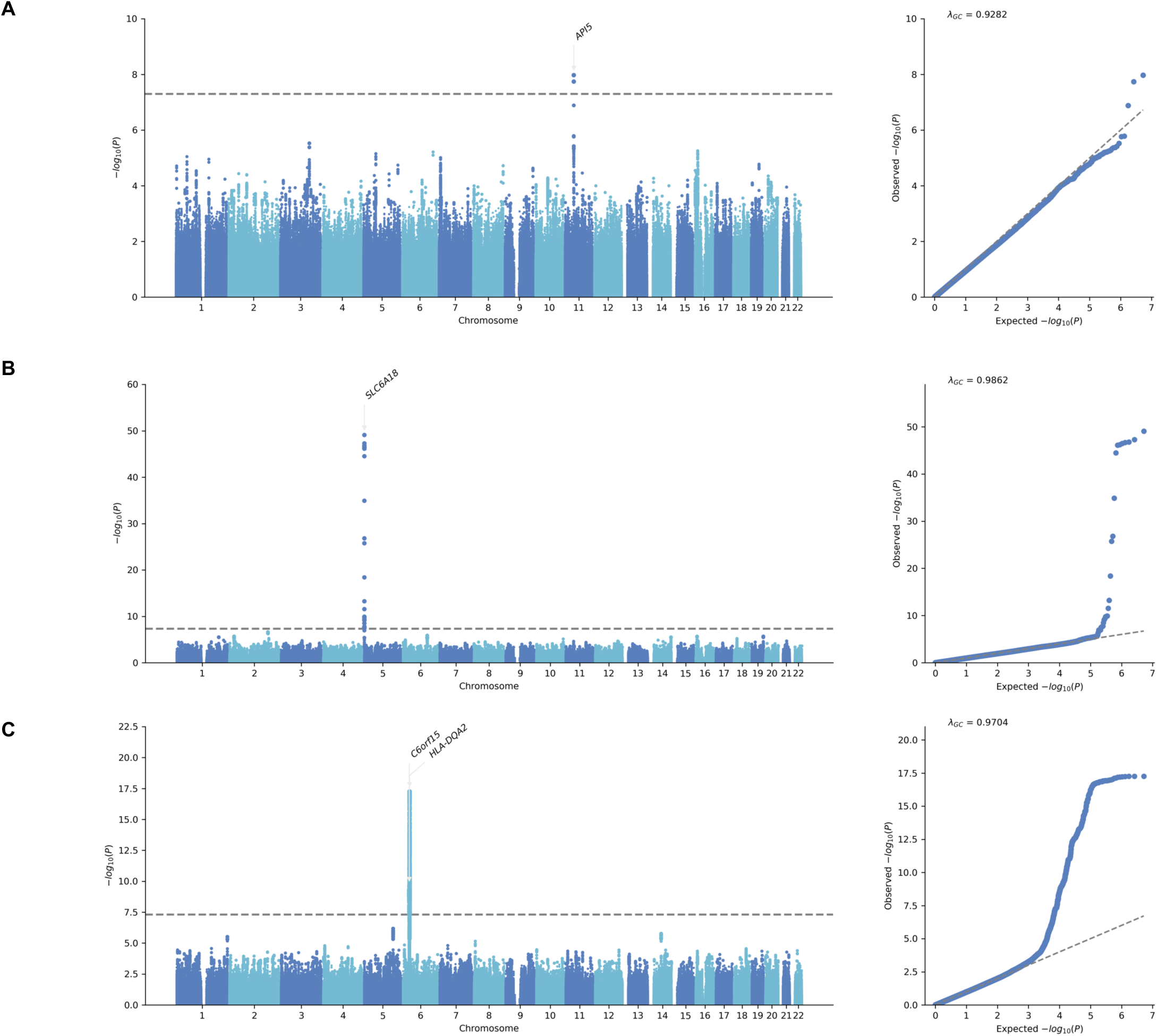
Manhattan plots of genome-wide association results for MDS, GIST, and ANSC. Manhattan plots of genome-wide association results for (**A**) myelodysplastic syndromes (MDS) (**B**) gastrointestinal stromal tumor (GIST), and (**C**) anal squamous cell carcinoma (ANSC). The y-axis shows –log₁₀ P-values, and the horizontal dashed line indicates the genome-wide significance threshold (P ≤ 5 × 10⁻⁸).

In germ cell tumors (1,590 cases), rs4690675-T near *GPM6A* was associated with increased risk in the three-cohort meta-analysis (OR = 2.11; *p* = 2.45×10^−8^). Effect directions were consistent across cohorts, with significant associations in IMPACT (OR = 1.89, *p* = 3.53×10^−5^) and FinnGen (OR = 4.43, *p* = 6.24×10^−5^) and concordant effect estimates in PROFILE (OR = 1.92, *p* = 0.48) (**Supplementary Table 3**).

In gastrointestinal stromal tumor (1,252 cases), we identified a highly significant association with rs62331325-A (OR = 1.91; *p* = 8.20×10⁻^−50^; **Figures 2B; 3A**). The variant lies within an intron of *SLC6A18* at 5p15.33, approximately 65 kb downstream of *TERT* and 88 kb downstream of *CLPTM1L*, two distinct oncogenes at this cancer susceptibility locus. The association replicated across all cohorts, with larger effect sizes observed in the clinically ascertained cohorts (PROFILE: OR = 2.59; IMPACT: OR = 2.15) compared to FinnGen (OR = 1.37). This finding implicates telomere biology in GIST susceptibility^27^.

In gastrointestinal neuroendocrine tumors (772 cases), rs3217810-T near *CCND2* reached genome-wide significance in the PROFILE–IMPACT meta-analysis (OR = 1.61; *p* = 1.61×10⁻⁹), with concordant effects across clinical cohorts but no evidence of association in FinnGen (**Supplementary Table 3**). This locus has previously been implicated in colorectal cancer GWAS, supporting biological plausibility in a related tissue type^28^.

In anal cancer (709 cases), rs17212748-T within the human leukocyte antigen (HLA) locus was associated with risk (OR = 1.58; *p* = 5.50×10^−18^; meta-analysis across PROFILE, IMPACT, and FinnGen; **Figures 2C, 4A**). The association was directionally consistent across cohorts, reaching genome-wide significance in the PROFILE GWAS and nominal significance in the independent IMPACT and FinnGen GWAS, with no evidence of heterogeneity in effect direction. The variant maps to the gene body of *HLA-DQA2*. Fine-mapping using imputed HLA alleles identified HLA-DQB1*03 as the only allele with posterior inclusion probability >0.8. This allele is in strong linkage disequilibrium with rs17212748-T (r^2^ = 0.927), supporting it as the likely causal variant (**Supplementary Figure 1**). A second independent locus, rs4371876-G (OR = 1.41; *p* = 2.54×10^−10^), located downstream of *C6orf15*, also reached genome-wide significance in the three-cohort meta-analysis, with consistent directions of effect across cohorts (**Supplementary Table 3**). This locus has been associated with other gastrointestinal malignancies^29^ but has not previously been reported in anal cancer, highlighting a novel extension of shared gastrointestinal susceptibility biology.

In non-melanoma skin cancer (29,285 cases), rs62336926-C near *MIR4277/MRPL36* demonstrated an association in the PROFILE–IMPACT meta-analysis (OR = 1.35; *p* = 9.92×10^−9^; **Supplementary Table 3**), whereas the effect was attenuated in the three-cohort meta-analysis (OR = 1.04; *p* = 5.27×10^−5^). This pattern is consistent with enhanced detection of disease-associated variants in clinically ascertained, pathology-confirmed cohorts compared to population biobanks.

In mesothelioma (1,340 cases), rs2736099-A at 5p15.33, located within the TERT gene body, was associated with increased risk (OR = 1.32; *p* = 6.26×10^−10^), with concordant effects across the three cohorts (**Supplementary Table 3**). Similarly, in hepatobiliary cancer (4,140 cases), rs10069690-C, also located within the TERT gene body, demonstrated a significant association (OR = 1.21; *p* = 2.38×10^−15^), with consistent effect sizes across the three cohorts. Together with the strong signal observed in GIST near *TERT*, these findings implicate the 5p15.33 region and telomerase regulation as a shared susceptibility mechanism across multiple rare malignancies arising in distinct tissue contexts. Notably, the lead variants for mesothelioma (rs2736099), hepatobiliary cancer (rs10069690), and GIST (rs62331325) are not in high linkage disequilibrium across ancestral populations (**Figure 5A**), indicating allelic heterogeneity with independent risk signals converging on the same locus. Consistent with this, cross-cancer analyses in UK Biobank and FinnGen reveal substantial pleiotropy at the *TERT* locus, with rs2736099 and rs10069690 showing genome-wide significant associations across multiple cancer phenotypes (**Figure 5B**). In contrast, rs62331325 appears to represent a more GIST-specific signal, with no prior genome-wide significant associations reported in population biobanks, highlighting it as a potentially novel variant at this locus.

### Germline–Somatic Interactions Reveal Subtype-Specific Effects in GIST

Given the unique availability of paired germline and somatic data in the clinical cohorts (PROFILE and IMPACT), we investigated, within each cancer type, whether lead GWAS risk variants identified in each cancer were associated with recurrent somatic alterations. For each cancer in which we observed a novel GWAS association, we tested associations between the lead risk SNP and somatic mutations occurring in at least 5% of patients, restricting analyses to recurrent and commonly observed somatic alterations with sufficient frequency to support stable estimation of germline–somatic associations.

In GIST, we examined the two most prevalent somatic drivers: *KIT* (altered in 75% of cases) and PDGFRA (6%), as well as quadruple-negative tumors (17%), defined as lacking mutations in known driver genes (see **Methods**). The A (risk) allele of the lead SNP rs62331325 was associated with a significantly increased likelihood of harboring a *KIT* mutation (OR = 2.21, *p* = 6.50×10^−4^) and a decreased likelihood of *PDGFRA* mutation (OR = 0.45, *p* = 0.017) in a meta-analysis of the PROFILE and IMPACT cohorts (**Figure 3B; Supplementary Table 5**). No association was observed with quadruple-negative GIST.

**Figure 3:**
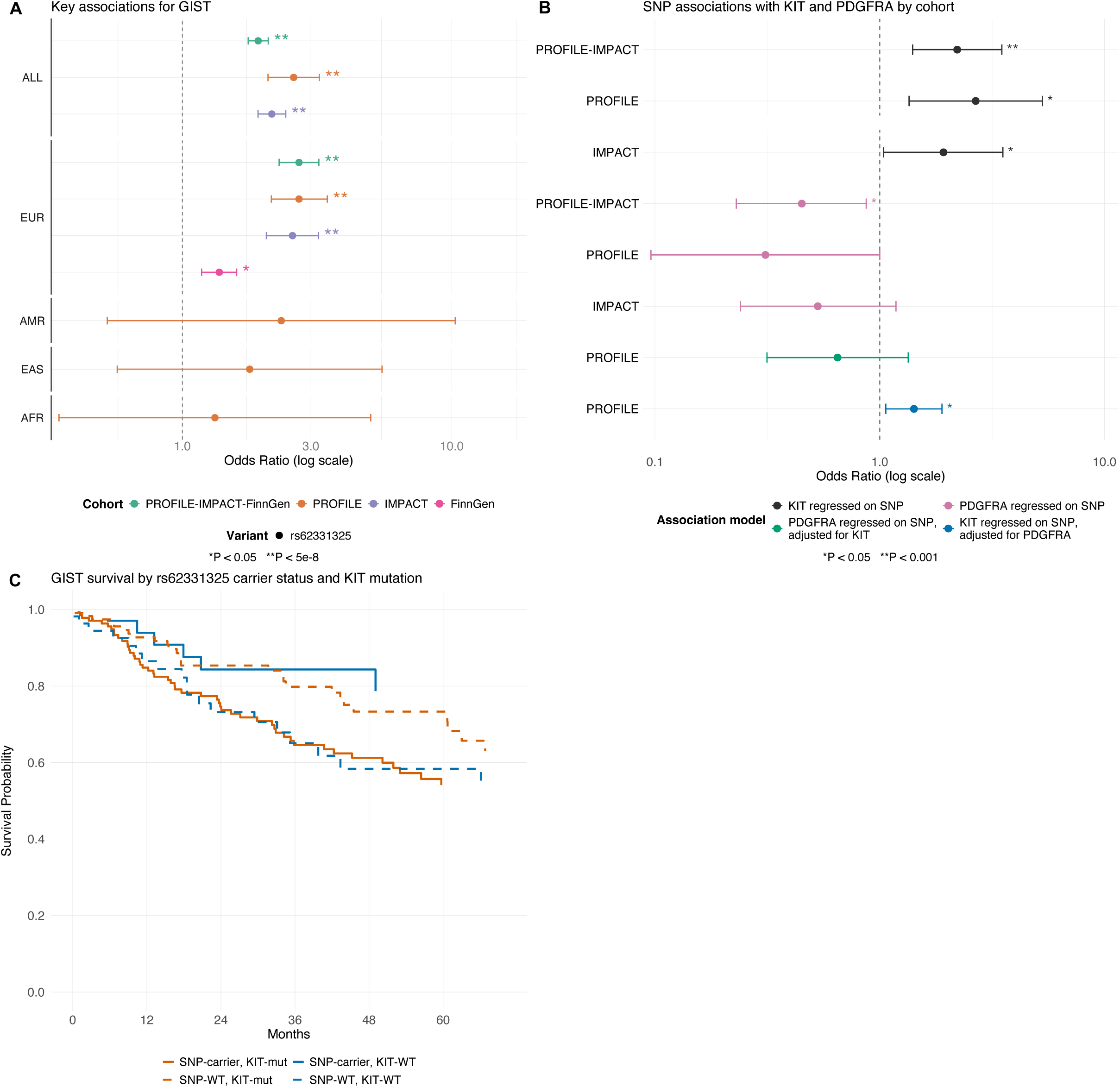
Lead germline SNPs GIST: GWAS Results, Somatic Mutation Association, and Survival Impact. (**A**) Forest plots showing genome-wide significant associations from the meta-analysis and cohort-specific analyses in GIST, stratified by ancestry. (**B**) Association of rs62331325, the lead variant from the GIST GWAS meta-analysis, with *KIT* or *PDGFRA* somatic mutation status in GIST patients. Effect sizes are shown across regression models (legend) and cohorts (PROFILE, IMPACT, and meta-analysis; y-axis). Ancestries not shown had insufficient case counts for ancestry-specific GWAS. Error bars represent 95% confidence intervals. (**C**) Kaplan–Meier survival analysis of patients with GIST, stratified by germline carrier status of rs62331325 (identified in the GIST GWAS) and by KIT somatic mutation status. Survival curves are shown for four groups: GWAS SNP carriers with KIT mutation, GWAS SNP carriers with KIT wild-type (WT), SNP non-carriers with KIT mutation, and SNP non-carriers with KIT WT. “SNP carrier” refers to carriers of SNP rs62331325-A. Solid lines represent SNP carriers; dashed lines represent SNP non-carriers.

Because both *KIT* and *PDGFRA* mutations were associated with the lead GIST SNP rs62331325-A, we performed conditional analyses to determine whether these signals reflected independent germline effects (see **Methods**). These analyses demonstrated that rs62331325-A was independently associated with the presence of *KIT* mutations (*p* = 0.017) (**Supplementary Table 6**). In contrast, the association with *PDGFRA* mutations (*p* = 0.24) was no longer significant after accounting for *KIT* status (**Supplementary Table 6**). Given the well-established mutual exclusivity between *KIT* and *PDGFRA* alterations in GIST, this pattern indicates that the primary effect of rs62331325-A is on *KIT*-mutant tumors, with the apparent association with *PDGFRA* arising indirectly from the increased prevalence of *KIT* mutations among risk allele carriers.

To determine whether the association between rs62331325-A and somatic driver mutations extends beyond GIST, we evaluated cancers in which *KIT* or *PDGFRA* somatic mutations occur at appreciable frequency (>1%). Specifically, we examined melanoma cases harboring *KIT* mutations (5% prevalence) and glioma cases with *PDGFRA* mutations (2% prevalence). rs62331325-A was not significantly associated with either somatic alteration in these cancers (**Supplementary Table 7**), indicating that the observed germline–somatic interaction is likely specific to the GIST context rather than reflecting a generalized relationship with *KIT* or *PDGFRA* mutations across tumor types.

Given the known prognostic relevance of *KIT* mutations in GIST^30^, we evaluated whether germline variation at rs62331325-A interacts with *KIT* mutation status to influence overall survival in the PROFILE cohort. In a Cox proportional hazards model, we observed a significant interaction between rs62331325-A carrier status and *KIT* mutation status (hazard ratio [HR] = 4.06, *p* = 0.015), suggesting that carriers of the risk allele with *KIT*-mutant GIST experience worse outcomes compared to patients without this combined risk profile (**Supplementary Table 8**). Replication analysis in the IMPACT cohort demonstrated an independent association between rs62331325-A and poorer survival after adjusting for *KIT* mutation status (HR = 1.85, *p* = 0.0047); however, the interaction between germline carrier status and *KIT* mutation was not statistically significant (**Supplementary Table 8**).

Collectively, these findings support a GIST-specific germline–somatic interaction in which rs62331325-A preferentially predisposes to *KIT*-mutant tumors and may influence clinical outcomes. The absence of similar associations in other *KIT*- or *PDGFRA*-mutant cancers underscores the context-dependent nature of this effect, while cross-cohort analyses suggest a shared germline risk signal with variability in the magnitude of survival interactions. Notably, effect directions for rs62331325-A were consistent across ancestry-stratified analyses, even in subgroups where limited sample sizes reduced statistical significance, supporting a shared underlying genetic effect across populations (**Figure 3A**).

Although associations with both KIT and PDGFRA mutations were observed, neither remained significant after correction for multiple testing across all somatic–germline analyses (FDR > 5%), consistent with more modest or context-dependent effects. Moreover, we did not observe comparable germline–somatic associations with recurrent alterations in the other rare cancers analyzed (**Supplementary Table 5**).

### Association of ANSC Risk Variant with HPV Susceptibility

Given the established link between ANSC and Human Papillomavirus (HPV) infection^31^ and the location of the novel risk variant in the Human Leukocyte Antigen (HLA) locus, we aimed to determine whether this lead SNP SNP in ANSC was mechanistically associated with HPV infection. We used data from the PROFILE cohort and derived HPV status from genomic sequencing reads mapped to viral genome references^32^. These estimates were highly concordant with clinical assays that were conducted in a subset of patients (see **Methods**), but enabled broader and less ascertained analysis of the entire cohort. As expected, HPV viral reads were significantly enriched (OR > 1) in cancers with known HPV etiology, including anal cancer (OR = 142.86, *p* = 1.85×10^−56^), cervical cancer (OR = 23.57, *p* = 9.39×10^−29^), and head and neck carcinoma (OR = 14.45, *p* = 9.39×10⁻^29^). In contrast HPV reads were rarely detected in cancers without established HPV involvement, including those arising in anatomically similar regions (**Supplementary Figure 2**).

We then focused on rs585305-T, which was the most statistically significant association in the PROFILE cohort with ANSC (**Supplementary Table 9**) and thus expected to best balance imputation accuracy and association with the outcome (though our findings were consistent when using rs17212748-T; **Supplementary Table 10**). rs585305-T was significantly associated with HPV viral read status in the overall population (OR = 1.36, *p* = 2.11×10⁻^4^), with a similar but more significant association in the European ancestry group (OR = 1.44, *p* = 3.19×10⁻^5^) after adjusting for cancer type (**Supplementary Table 10**). Because rs585305-T is both an HPV risk–increasing allele and an ANSC risk–increasing allele, the variant may elevate ANSC risk through increased susceptibility to HPV infection, which typically precedes the development of HPV-related cancers^31^.

We extended our analysis of this variant to additional HPV-associated cancers, defined as those with more than 50 HPV-positive cases and an HPV prevalence of at least 1%, to evaluate whether the association was specific to ANSC risk or more broad. rs585305-T showed significant association with HPV status in cervical cancer (OR = 2.00, *p* = 2.20×10⁻^4^) and in ANSC (OR = 1.68, *p* = 0.038), respectively, but was not significant in head and neck carcinoma (OR = 1.15, *p* = 0.13) (**Figure 4B**, **Supplementary Table 11**). While these associations do not reach genome-wide significance, they suggest that this ANSC-linked locus may also contribute to HPV infection in other HPV-driven cancers.

**Figure 4:**
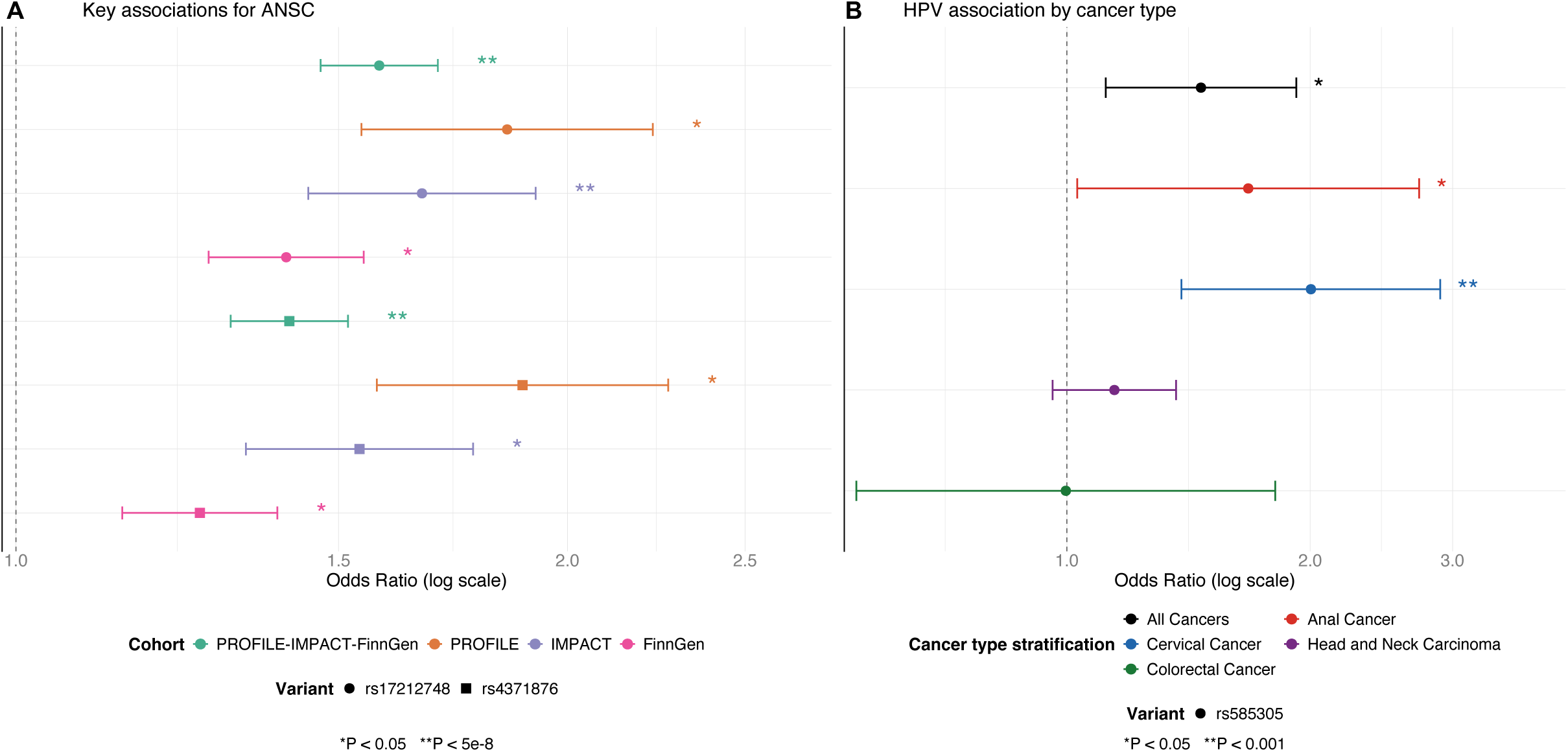
Lead germline risk variants for ANSC and association with HPV. (**A**) Forest plot of the genome-wide significant associations at the lead meta-analysis variants rs17212748-T and rs4371876-G for anal squamous cell carcinoma (ANSC), showing effect estimates across cohorts using analyses performed in all individuals (not stratified by ancestry). Error bars represent 95% confidence intervals. (**B**) Association of rs585305-T, the lead variant from the ANSC GWAS in the PROFILE cohort, with HPV status across HPV-related cancer types (y-axis; legend), based on analyses performed in all individuals (not stratified by ancestry) within the PROFILE cohort.

**Figure 5:**
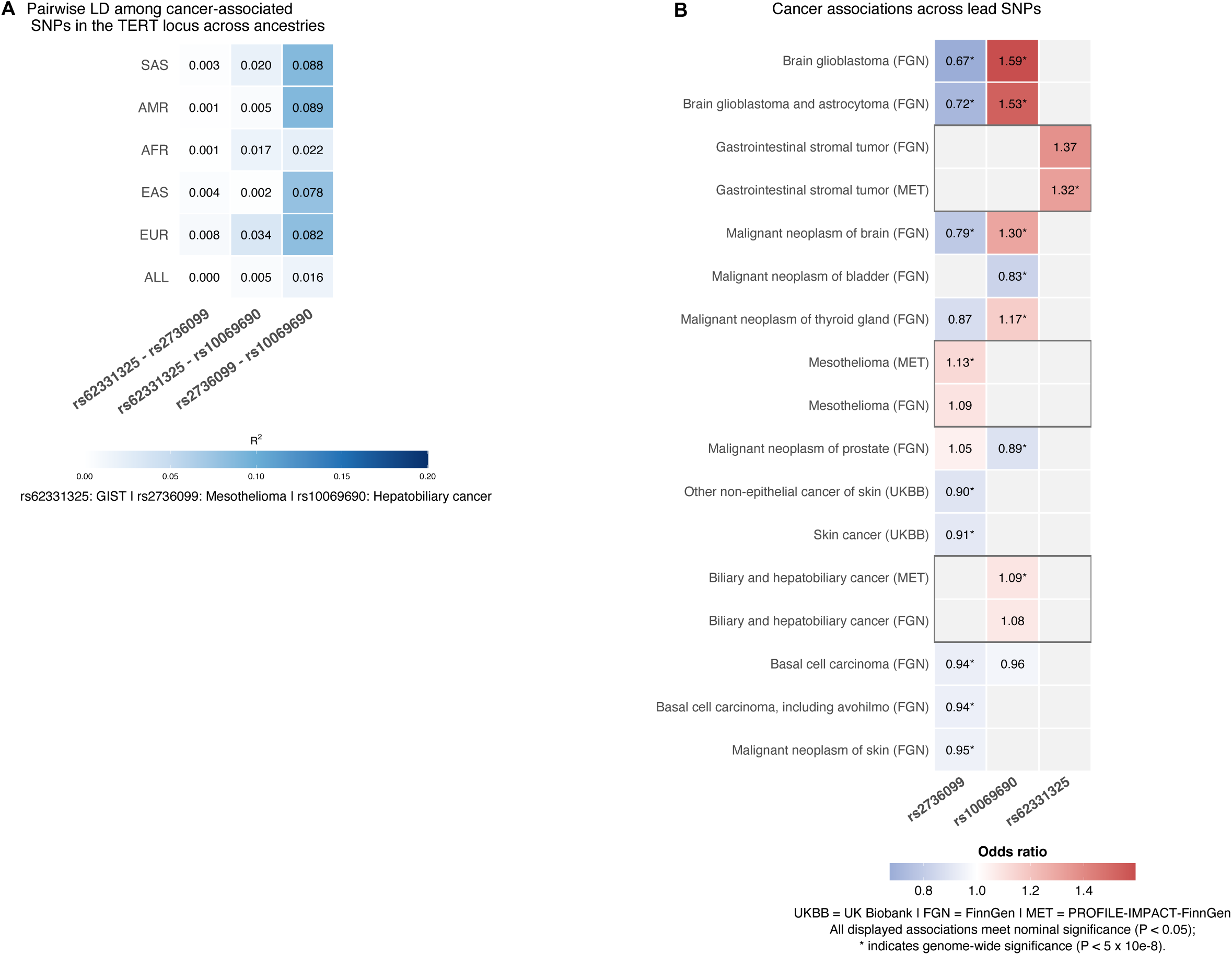
Linkage disequilibrium and cross-cancer association landscape of rare cancer GWAS signals at the TERT locus. (**A**) Pairwise linkage disequilibrium ($^!^ (between three rare cancer GWAS lead variants at the TERT locus, rs62331325-A (gastrointestinal stromal tumor), rs2736099-A (mesothelioma), and rs10069690-C (hepatobiliary cancer), across ancestral populations from the 1000 Genomes Project reference panel, computed using LDlink. (**B**) Cross-cancer association profiles of each lead variant at the TERT locus across UK Biobank, FinnGen, and the PROFILE-IMPACT-FinnGen meta-analysis conducted in this study. Phenotypes shown are restricted to those with at least one genome-wide significant association. Tile color represents effect size as odds ratio. Asterisks denote genome-wide significance (% < 5×10^−8^). All plotted odds ratios correspond to at least nominally significant associations (%<0.05); gray tiles indicate associations that do not reach nominal significance. Boxes highlight the primary rare cancer phenotype–SNP associations identified in this study, whereas unboxed phenotypes represent additional cancer phenotypes from population biobanks (UK Biobank and FinnGen) that are also significantly associated with these variants.

### API5-Linked Apoptotic Mechanism in MDS

The variant implicated in myelodysplastic syndromes (MDS), rs76521016-C, is located upstream of *API5*, a gene encoding apoptosis inhibitor 5, a well-characterized regulator of programmed cell death^26^. *API5* suppresses apoptosis and has been implicated in the survival and persistence of abnormal cells across multiple malignancies, including hematologic disorders. Dysregulation of *API5* expression has been associated with enhanced cellular survival and resistance to apoptotic cues, providing a biologically plausible mechanism for altered hematopoietic cell dynamics.

Motivated by this functional context, we evaluated whether rs76521016-C was associated with routine clinical hematologic measurements obtained proximal to the tumor biopsy date. We tested associations with six laboratory traits: absolute neutrophil count, white blood cell count, hemoglobin, hematocrit, red blood cell count, and platelet count (see **Methods**). Among these traits, rs76521016-C was significantly associated with absolute neutrophil count (β = 77.72, *p* = 0.0046), with each additional copy of the effect allele corresponding to an increase in neutrophil count (**Supplementary Table 12)**. The observed effect size is consistent with a model in which the MDS-associated GWAS variant influences *API5* regulation, potentially attenuating apoptosis and thereby prolonging neutrophil survival in carriers of the effect allele. No significant associations were observed for the remaining laboratory traits or hematologic cell types.

This association is biologically consistent with the known role of apoptosis in regulating neutrophil homeostasis. Neutrophils are the most short-lived circulating hematopoietic cells and rely heavily on tightly regulated apoptotic pathways to maintain appropriate turnover^33^. In contrast, erythroid cells and platelets have longer lifespans or are buffered by precursor pools, potentially rendering them less sensitive to modest perturbations in apoptotic signaling^34^. The selective association with neutrophil counts therefore supports a model in which variation near *API5* influences myeloid cell survival, rather than broadly affecting hematopoiesis.

Together, these findings provide functional support for the genetic association observed in myelodysplastic syndromes and suggest that rs76521016-C may contribute to disease biology through modulation of apoptosis-dependent neutrophil survival.

### TWAS and RWAS Provide Functional Resolution of GWAS Loci

To investigate the functional relevance of the GWAS loci identified, we performed transcriptome-wide association (TWAS) and regulome-wide association (RWAS) analyses for each cancer, focusing on regions surrounding the lead variants. TWAS identifies nearby genes whose genetically predicted expression is associated with GWAS loci, whereas RWAS tests for associations with genetically predicted regulatory activity. Across cancers, these complementary approaches revealed both recurrent cross-cancer convergence at established susceptibility loci and locus-specific complexity in regions characterized by dense linkage disequilibrium (LD).

The most prominent convergence occurred at the *TERT*–*CLPTM1L* locus on chromosome 5p15.33, a well-established pan-cancer susceptibility region^35–38^. In hepatobiliary cancer, TWAS identified a significant association with *TERT* (Bonferroni-adjusted *p* = 6.67×10⁻⁵; **Supplementary Table 13**). Consistently, RWAS detected two highly significant regulatory elements near *TERT*, located at chr5:1,345,008–1,345,509 (50 kb upstream of *TERT*) and chr5:1,293,863–1,294,364 (overlapping the *TERT* transcription start site), with Bonferroni-adjusted p-values of 8.22×10^−9^ and 4.76×10^−15^, respectively (**Supplementary Table 13**). Mesothelioma showed a similar pattern, with TWAS implicating *TERT* (Bonferroni-adjusted *p* = 0.0043) and RWAS identifying a significant peak at chr5:1,345,008–1,345,509 (50 kb upstream of *TERT*; Bonferroni-adjusted *p* = 0.0064; **Supplementary Table 13**). Notably, the same upstream regulatory element at 5p15.33 (chr5:1,345,008–1,345,509) was significantly associated in both mesothelioma and hepatobiliary cancer, indicating convergence on a shared regulatory region. However, the lead GWAS variants for these cancers are not in high linkage disequilibrium, demonstrating that these represent genetically independent association signals (**Figure 5A**). Together, these findings suggest that distinct risk variants may converge on a common regulatory element influencing *TERT* expression across tumor types.

Associations with GIST implicated additional mechanisms in the 5p15.33 region. Five genes within 1 Mb of the lead GWAS locus rs62331325 reached Bonferroni significance in TWAS, including *RP11-43F13.3* (*p* = 2.75×10^−5^), *CTD-3080P12.3* (*p* = 1.74×10^−3^), *NKD2* (*p* = 6.68×10^−3^), *ZDHHC11B* (*p* = 8.70×10^−3^), and *CLPTM1L* (*p* = 1.80×10^−2^; **Supplementary Table 13**). RWAS identified three significant regulatory elements, all localizing near *SLC6A18*, the gene containing the lead intronic variant. Two peaks located 72 kb and 52 kb upstream of the *SLC6A18* transcription start site (chr5:1,153,486–1,153,987, *p* = 1.78×10^−3^; and chr5:1,173,133–1,173,634, *p* = 3.76×10^−3^, respectively) and a third peak located 20 kb downstream of *SLC6A18* and 8 kb downstream of *TERT* (chr5:1,245,038–1,245,539, *p* = 1.54×10^−2^) suggest that altered regulatory activity across the broader locus may contribute to GIST susceptibility (**Supplementary Table 13**). Several implicated genes at this locus have established cancer relevance, including *NKD2*, a negative regulator of Wnt signaling^39^; *ZDHHC11B*, reported as a tumor suppressor^40^; and *CLPTM1L*, linked to anti-apoptotic processes^41^. Importantly, the TWAS signal for *CLPTM1L*, together with regulatory enrichment across the broader 5p15.33 landscape, suggests that *CLPTM1L* may represent an additional—and potentially primary—effector gene at this canonical susceptibility locus, alongside *TERT*. Together, these findings support a model in which regulatory variation at the *CLPTM1L*–*TERT* locus influences rare cancer susceptibility through effects on telomere maintenance (via *TERT*) and apoptosis or survival-related pathways (via *CLPTM1L*).

Several cancers also exhibited locus-specific gene-level associations that closely aligned with nearby GWAS variants. In gastrointestinal neuroendocrine tumors (GI-NET), TWAS identified *CCND2* as significantly associated with the lead GWAS variant rs3217810 (Bonferroni-adjusted *p* = 1.61×10^−9^; **Supplementary Table 13**). Given that *CCND2* encodes cyclin D2, a central regulator of cell-cycle progression, the concordance between the GWAS locus and TWAS association suggests that altered *CCND2* expression may directly mediate inherited susceptibility. In myelodysplastic syndromes (MDS), TWAS identified *TTC17* as significantly associated (Bonferroni-adjusted *p* = 0.037), despite the lead GWAS signal mapping upstream of *API5*, indicating regulatory complexity and potential alternative target genes within the locus. *TTC17* has been implicated in cancer through its role in Golgi organization and vesicular trafficking, supporting its candidacy as a functionally relevant gene at this locus^42^.

ANSC also demonstrated a concentration of associations within the highly polymorphic HLA region, where 24 genes reached Bonferroni significance in TWAS and 13 genomic regions were significant in RWAS (**Supplementary Table 13**). The TWAS-implicated genes included *AGER*, *AGPAT1*, *BRD2*, *BTNL2*, *HCG23*, *HLA-DPA2*, *HLA-DPB1*, *HLA-DQA1*, *HLA-DQA2*, *HLA-DQB1*, *HLA-DQB1-AS1*, *HLA-DQB2*, *HLA-DRA*, *HLA-DRB1*, *HLA-DRB5*, *HLA-DRB6*, *NOTCH4*, *PBX2*, *PPP1R2P1*, *RNF5*, *RPL32P1*, *TAP2*, *XXbac*-*BPG154L12.4*, and *XXbac*-*BPG254F23.7*. The extreme LD structure, high gene density, and extensive polymorphism characteristic of this region likely complicate accurate imputation of gene expression and regulatory activity.

Conditional analysis of the RWAS signal in hepatobiliary cancer (rs10069690) showed substantial attenuation (from *p* = 1.0×10^−15^ to *p* = 3.6×10^−4^ after conditioning on the lead GWAS variant), consistent with the RWAS association being largely explained by the primary GWAS locus. Across loci, most TWAS and RWAS signals did not meet the threshold for follow-up conditional analysis.

Overall, TWAS and RWAS analyses provide functional resolution for multiple GWAS loci, revealing both recurrent cross-cancer convergence—most prominently at the *TERT*–*CLPTM1L* locus—and locus-specific complexity in regions such as HLA. The alignment of gene-level and regulatory-level associations strengthens the biological plausibility of implicated targets while highlighting the complex local genetic architecture underlying several susceptibility loci.

## Discussion

In this study, we performed a meta-analysis of germline genetic variation across 20 rare cancer types by integrating imputed germline data from two large clinically curated cancer sequencing cohorts with the FinnGen population biobank. By leveraging cancer-specific cohorts in which germline genotypes were imputed from routine tumor panel sequencing, our framework increased case counts for each cancer and enabled a systematic evaluation of rare cancer susceptibility across diverse malignancies, rather than focusing on individual tumor types in isolation. This approach led to the discovery of both cancer-specific and cross-cancer susceptibility loci that were reproducible across independent cohorts. In total, we identified nine novel genome-wide significant associations with moderate to large effect sizes. These findings demonstrate that even cancers traditionally considered underpowered for GWAS can yield robust germline discoveries when case counts are substantially expanded through clinically ascertained cancer cohorts.

Across the rare cancers analyzed, effect sizes were frequently larger than those typically observed for common variants in more common cancers, suggesting that rare cancers may be influenced by higher-impact common germline variants. The strongest associations were observed for gastrointestinal stromal tumor (GIST), myelodysplastic syndromes (MDS), germ cell tumor, and anal cancer (ANSC), with odds ratios ranging from 1.9-2.6. These magnitudes are notable for common variants and rank among the largest reported for cancer-related traits in the GWAS catalog. For example, the GIST association (OR ≈ 2.6) exceeds that reported for any variant with allele frequency >5% for solid cancers in the UK Biobank^43^. Consistent with these large effect sizes, population attributable risk (PAR) estimates were also substantial^44,45^. In the PROFILE cohort (European ancestry), PAR was estimated at approximately 23% for ANSC and 30% for GIST, indicating that these loci contribute meaningfully to disease burden at the population level (see **Methods**). Together, these findings suggest that, unlike many common cancers where risk is highly polygenic and individually modest, certain rare cancers may be driven in part by common variants of comparatively large effect.

Across the remaining cancers with genome-wide significant associations, we observed signals in mesothelioma, hepatobiliary cancer, and gastrointestinal neuroendocrine tumors. Several of these signals converged on shared biological processes—most prominently telomere maintenance at the *TERT–CLPTM1L* locus across mesothelioma, hepatobiliary cancer and GISTr—suggesting that distinct rare malignancies may share partially overlapping germline architectures despite arising in different tissues. Although these variants map to the same locus, they are not in strong linkage disequilibrium, indicating that they represent multiple independent risk signals converging on a shared regulatory region near TERT. Consistent with this interpretation, two of the variants have been implicated in distinct cancer phenotypes in prior population-based studies, supporting pleiotropic but independent effects, whereas the GIST-associated variant appears to represent a novel, cancer-specific signal at this locus. Other associations appeared more cancer-specific, including the HLA locus in ANSC, implicating immune-mediated susceptibility in HPV-driven tumorigenesis; the signal near *API5* in MDS, consistent with apoptosis dysregulation in hematopoietic cells; and the association near *CCND2* in gastrointestinal neuroendocrine tumors, pointing to altered cell-cycle control. Notably, none of the associations reached genome-wide significance in FinnGen alone, underscoring the value of incorporating clinically sequenced cancer cohorts to increase effective case counts and enhance discovery power.

The availability of detailed clinical and molecular data in the PROFILE cohort, features not typically accessible in population biobank GWAS, enabled more granular mechanistic follow-up of selected loci. In GIST, the rs62331325-A risk allele was associated with increased likelihood of *KIT*-mutant tumors and with poorer survival among carriers with *KIT* mutations, suggesting that germline variation may influence both tumor subtype and clinical trajectory. Conditional analyses indicated that the inverse association with *PDGFRA* mutations was likely mediated through mutual exclusivity with *KIT*. Because *KIT*-mutant and *PDGFRA*-mutant GISTs arise preferentially in different anatomical sites, rs62331325-A may exert tissue-context–dependent effects on tumor initiation or on selection of driver mutations^46^.

In ANSC, the lead variant rs17212748-T lies within the HLA locus, a region central to antigen presentation and adaptive immune regulation. This localization suggests that inherited variation may influence host immune responses to oncogenic viral infection. Consistent with this hypothesis, the risk allele was associated with HPV viral read status not only in ANSC but also in other HPV-driven cancers. These findings support a model in which germline immune variation increases susceptibility to persistent or higher-burden HPV infection, thereby elevating downstream cancer risk. More broadly, this locus illustrates how inherited differences in immune surveillance may shape infection-mediated carcinogenesis.

In MDS, the rs76521016-C risk allele, located upstream of *API5*, a regulator of apoptosis, was associated with increased neutrophil counts, supporting a role in apoptosis-dependent myeloid cell survival. Notably, this association was selective for neutrophils and was not observed for red blood cell or platelet counts, suggesting a lineage-specific effect rather than broad disruption of hematopoiesis. Given the short lifespan of neutrophils and their reliance on tightly regulated apoptotic turnover, this pattern is consistent with a model in which altered *API5* activity prolongs neutrophil survival, linking germline variation to a measurable intermediate cellular phenotype in MDS.

TWAS and RWAS analyses provided functional support for several GWAS loci, most prominently highlighting recurrent cross-cancer convergence at the *TERT–CLPTM1L* locus (5p15.33). Across GIST, hepatobiliary cancer, and mesothelioma, gene-level and regulatory-level associations implicated *TERT* and nearby genes, suggesting coordinated effects on telomere biology. *TERT* is notably upregulated in GIST and mesothelioma despite the absence of frequent somatic promoter mutations^47,48^. Conversely, in hepatobiliary cancer, *TERT* activation is commonly driven by recurrent somatic promoter mutations^49,50^.

Despite these insights, our study has several limitations. The small sample sizes for non-European ancestry subpopulations likely restricted our ability to detect associations that are specific to those groups. Additionally, the use of imputed genotypes, while expanding genomic coverage, introduces a layer of uncertainty, particularly in genetically complex regions like the

HLA locus. These factors could potentially impact the precision of our estimated effect-sizes, though we expect imputation error to generally lead to underestimation. Additionally, a large fraction of our cases came from two academic medical centers via routine tumor sequencing. This selected patient group may not reflect the general patient population and risk profile. Interestingly, the effect sizes we observed in these two cohorts were generally higher than those observed in FinnGen, a less ascertained cohort that includes non-cancer controls. This suggests that germline associations may appear stronger in cancer cohorts for several reasons: (1) phenotype definitions are more precise in cancer center datasets than in population biobanks, (2) patients with more aggressive tumors are more likely to undergo biopsy and next-generation sequencing, enriching cancer cohorts for more extreme phenotypes, and (3) the use of population-based rather than fully matched controls in FinnGen may attenuate effect sizes.

This study provides evidence that rare cancers harbor reproducible germline susceptibility loci and demonstrates that discovery in these malignancies is both feasible and informative when case ascertainment is expanded through clinically sequenced cancer cohorts. By systematically analyzing 20 rare tumor types, we identified multiple novel common-variant susceptibility loci and uncovered both shared biological pathways and cancer-specific mechanisms. Our results show that integrating clinically ascertained sequencing cohorts with population biobanks substantially enhances discovery power, enabling identification of high-confidence loci that would be missed in population cohorts alone and facilitating downstream mechanistic interpretation through linked somatic, viral, and clinical data. Notably, even cancers traditionally viewed as predominantly environmentally driven, such as HPV-associated anal cancer, exhibit strong germline genetic modifiers, underscoring the interplay between inherited susceptibility and external exposures. Together, these findings establish a scalable framework for elucidating the inherited architecture of rare malignancies and provide a foundation for future studies aimed at risk stratification, mechanistic studies, and therapeutic investigation across diverse tumor types.

## Methods

### Study populations, sequencing platforms and phenotype ascertainment

We analyzed two clinically sequenced cancer cohorts: the Dana-Farber PROFILE cohort (35,260 tumors spanning >62 cancer types) and the Memorial Sloan Kettering MSK-IMPACT cohort (54,945 patients). Tumors were sequenced using hybrid capture–based targeted next-generation sequencing panels: OncoPanel in PROFILE (275–447 cancer-associated genes across three versions, coding exons only) and IMPACT in MSK-IMPACT (341–505 genes across four versions, including coding exons and selected introns capturing clinically actionable alterations).

In both cohorts, tumor sequencing enabled systematic profiling of recurrent somatic alterations, including oncogenic driver mutations across cancer types. Germline genotypes were imputed from tumor-only sequencing data using STITCH with the 1000 Genomes Project reference panel. This approach has been previously validated, demonstrating a mean genotype correlation of 0.86 with matched germline SNP array data after quality control^13^.

In addition to genomic data, the clinical sequencing cohorts were linked to institutional clinical records. PROFILE included routinely collected laboratory measurements, curated structured data and unstructured clinical documentation. Both clinical cohorts included longitudinal survival data.

FinnGen is a population-based biobank comprising 379,154 participants with genotype data linked to nationwide health registry records. Genotyping was performed using customized Illumina arrays, and imputation was conducted using a Finnish population–specific whole-genome sequencing reference panel. FinnGen summary statistics were used for the following cancer endpoints: anal cancer (finngen_R12_C3_ANUS_ANALCANAL), hepatobiliary cancer (finngen_R12_C3_BILIARY_GALLBLADDER), gastrointestinal stromal tumor (GIST; finngen_R12_C3_GASTRINT_STROM), mesothelioma (finngen_R12_C3_MESOTHELIOMA), non-melanoma skin cancer (NMSC; finngen_R12_C3_OTHER_SKIN), and germ cell tumor (finngen_R12_C3_TESTIS).

In PROFILE and MSK-IMPACT, cancer phenotypes were defined using curated clinical and histopathologic annotations. Individuals diagnosed with a given cancer were classified as cases for that phenotype; all other sequenced individuals were treated as controls. In FinnGen, cancer phenotypes were defined using registry-based diagnostic codes; cases were individuals with a recorded diagnosis of the cancer of interest and controls were participants without any cancer diagnosis.

### Genome-Wide Association Analysis and Fine Mapping HLA Locus

Genome-wide association analyses were performed using REGENIE^24^ (v3.0.2), a ridge regression–based framework optimized for large-scale genetic association studies. Genotypes were imputed prior to association testing, and SNP dosages (expected allele counts) were used as input for association analyses in REGENIE. REGENIE operates in two stages: in stage 1, a whole-genome ridge regression model is fit to estimate the polygenic component of the trait; in stage 2, single-variant association tests are conducted using residuals from the stage 1 model. All GWAS models included sequencing panel version, sex, age, and the first five principal components of genetic ancestry (PC1–PC5) as covariates.

For downstream analyses, including meta-analysis, transcriptome-wide association studies (TWAS), and regulatory element–wide association studies (RWAS), summary statistics were restricted to variants present in all three cohorts (DFCI-PROFILE, MSK-IMPACT, and FinnGen) to ensure harmonized variant sets and balanced contribution across datasets.

Meta-analyses across cohorts were conducted using REMETA^25^, combining study-specific effect estimates using inverse-variance weighting. Ancestry-stratified GWAS were performed within continental ancestry groups using REGENIE, followed by meta-analysis with REMETA.

Fine-mapping of imputed HLA alleles was conducted in the PROFILE cohort to refine associations at the ANSC risk locus. Two-digit and four-digit HLA allele dosages were filtered based on imputation quality (R² > 0.75), retaining high-confidence alleles for analysis. Allele-specific associations were tested using logistic regression, adjusting for age, sex, sequencing panel version, and ancestry principal components. Analyses were performed in both the full cohort and the European ancestry subset. Marginal z-scores and the corresponding allele-level linkage disequilibrium (LD) matrix were used as input to the SuSiE fine-mapping algorithm^51^ (susie_rss) to estimate posterior inclusion probabilities (PIPs) and construct 95% credible sets.

### Germline–Somatic Interaction Analyses

In the PROFILE and MSK-IMPACT cohorts, REGENIE^24^ was used to test associations between germline variants and somatic mutation status, with mutation calls derived from tumor sequencing data.

For analyses of rs62331325 in gastrointestinal stromal tumor (GIST), quadruple-negative cases^52^ were defined as tumors lacking mutations in *KIT, PDGFRA, SDHA, SDHAF2, SDHB, SDHC, SDHD, KRAS, NRAS, HRAS, BRAF, RAF1, ARAF, RASA1, NF1, SOS1, MAP2K1, MAP2K2,* and *MAPK1*.

Conditional logistic regression analyses were performed to evaluate the relationship between rs62331325 and *KIT* and *PDGFRA* mutation status. Two complementary models were fit: (1) *KIT* mutation status as the outcome, with SNP dosage and *PDGFRA* mutation status as predictors (*KIT* ∼ SNP + *PDGFRA*), and (2) *PDGFRA* mutation status as the outcome, with SNP dosage and *KIT* mutation status as predictors (*PDGFRA* ∼ SNP + *KIT*). A joint model was additionally fit with SNP dosage as the dependent variable and both *KIT* and *PDGFRA* mutation status as predictors (*SNP* ∼ *KIT* + *PDGFRA*).

### HPV-Related Analyses in ANSC

Viral-based HPV status was determined in the PROFILE cohort using a previously validated computational pipeline^32^ for detection of DNA viruses in tumor sequencing data. Sequencing reads not aligning to the human reference genome were digitally subtracted and analyzed for viral content. HPV-positive cases were defined as samples with more than two viral reads aligning to any known HPV strain (**Supplementary Table 14**). This threshold identified 527 HPV-positive tumors. HPV16 was the most frequently detected strain (n = 467), followed by HPV18 (n = 25). Additional HPV types were observed at lower frequencies, including high-risk (e.g., HPV53) and low-risk strains (e.g., HPV6b, HPV10, HPV4). All detected HPV types were included in downstream analyses to maximize sensitivity for identifying viral associations.

Clinical HPV status was independently ascertained from electronic health record (EHR) data using a locally deployed Llama 3 8B large language model. All patients across cancer types at Dana-Farber Cancer Institute with available clinical documentation, including progress notes, pathology reports, and imaging reports from 2010 onward, were identified. Among these, 11,788 patients had at least one clinical note containing the term “HPV.”

Unstructured clinical text was processed on a per-note basis using a standardized prompt to extract structured HPV-related information. The model was configured to extract the following attributes: (1) HPV status, classified as positive, negative, or not mentioned; (2) HPV subtype, such as HPV16 or HPV18; (3) testing modality, including PCR or DNA sequencing; and (4) additional relevant annotations, such as p16 status or treatment eligibility.

Patient-level HPV status was assigned using a predefined rule-based aggregation strategy. Patients were classified as HPV-positive if any note documented positive HPV status with a specified subtype. Patients were classified as “not mentioned” if all notes lacked documentation of HPV status. All remaining patients were classified as HPV-negative. Application of this framework identified 2,172 HPV-positive patients, 7,837 HPV-negative patients, and 1,779 patients without documented HPV status.

### Functional Follow-Up Analyses: TWAS and RWAS

Transcriptome-wide association studies (TWAS) and regulome-wide association studies (RWAS) were conducted to evaluate whether genetically predicted gene expression or chromatin accessibility was associated with rare cancer risk. For each gene or regulatory element, predictive models of molecular activity were trained using common germline variants located within the cis-regulatory window of the feature. Only features demonstrating significant cis-heritability and adequate cross-validated prediction performance were retained for downstream analysis. For each lead GWAS locus, analyses were restricted to genes or regulatory elements located within ±500 kb of the index SNP to prioritize proximal cis-regulatory effects.

Prediction weights from these models were applied to the meta-analyzed germline rare cancer GWAS summary statistics generated in this study to test for associations between genetically predicted gene expression or regulatory element activity and rare cancer phenotypes. Analyses were performed using the FUSION framework^53^, with standard quality control filters applied to GWAS summary statistics, including restriction to HapMap3 variants to capture the majority of common SNP heritability and filtering based on imputation accuracy. Multiple testing correction was applied separately within TWAS and RWAS for each locus based on the number of genes or regulatory elements evaluated within the ±500 kb window.

Gene expression prediction models were derived from RNA sequencing data generated by TCGA, GTEx, and other eQTL reference panels. Chromatin accessibility models were trained using TCGA ATAC-seq data^53,54^. Each reported association corresponds to a gene or regulatory element evaluated within a specific tissue- or cohort-derived reference model. Because cis-QTL effects are frequently shared across tissues, associations were often observed across multiple tissue-specific models for the same molecular feature.

### Association of rs76521016 with Clinical Hematologic Traits in MDS

The association between the myelodysplastic syndromes (MDS)–associated variant rs76521016 and routine hematologic measurements was evaluated in the PROFILE cohort. Individuals were eligible for analysis if genotype data were available and at least one qualifying routine blood test measurement had been recorded.

For each laboratory test, the measurement obtained closest to the tumor biopsy date was selected. Six hematologic traits were analyzed: absolute neutrophil count, white blood cell count, hemoglobin, hematocrit, red blood cell count, and platelet count. Each trait was evaluated separately using the standardized laboratory value as the dependent variable.

Associations between rs76521016 and each hematologic trait were tested using linear regression under an additive genetic model, with genotype coded as effect-allele dosage. Models were adjusted for age at biopsy, sex, sequencing panel version, and the first five principal components of genetic ancestry.

### Population Attributable Risk Estimation

Population attributable risk (PAR) was estimated for a given variant using the formula PAR = [P(OR - 1)] / [P(OR - 1) + 1], where P is the allele frequency and OR is the odds ratio. Estimates were calculated using PROFILE cohort effect sizes and allele frequencies from gnomAD for European ancestry.

### Software and Tools

● GWAS analyses: REGENIE^24^ v3.0.2
● Meta-analysis: REMETA^25^
● Genotype imputation: HRC/1000 Genomes; GLIMPSE v1.1.1, STITCH
● TWAS/RWAS: FUSION
● PCA and ancestry inference: PLINK 2.0
● Viral detection: digital subtraction pipeline^32^
● EHR text mining: Meta-Llama-3-8B

All statistical analyses were conducted using Python and R. Figures were generated using ggplot2 and matplotlib.

## Data Availability

This study used data from the PROFILE and IMPACT cohorts. These datasets contain individual-level clinical and genomic information and are not publicly available due to privacy and institutional restrictions. However, summary statistics generated in this study from PROFILE and IMPACT will be made available.

We also used publicly available FinnGen GWAS summary statistics.

## Inclusion & Ethics Statement

Participants were recruited from hospital-based cancer cohorts (DFCI-PROFILE and MSK-IMPACT) and the population-based FinnGen biobank, with all individuals providing informed consent. Ethics approval was obtained from the Dana-Farber/Harvard Cancer Center IRB, the Memorial Sloan Kettering Cancer Center IRB, and the Coordinating Ethics Committee of the Hospital District of Helsinki and Uusimaa.

## Data Availability

FinnGen summary statistics are publicly available online. Summary statistics for the DFCI-PROFILE and MSK-IMPACT cohorts will be made available upon publication of the manuscript.

---

**Supplementary Figure 1:**
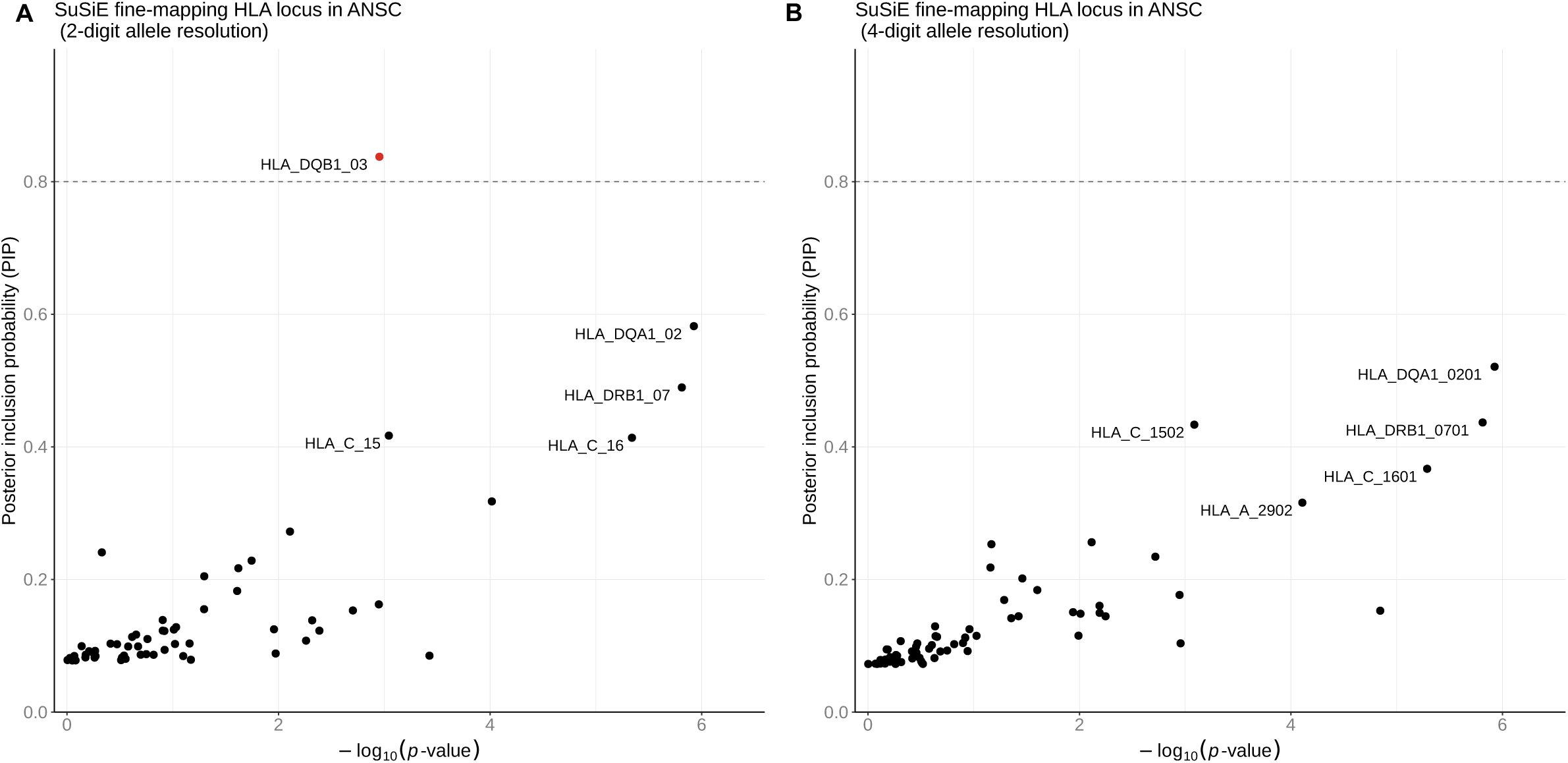
Fine Mapping of HLA at ANSC Locus. Each point represents an HLA allele group [2-digit] (**A**) or specific allele [4-digit] (**B**), with the x-axis showing the GWAS association strength (−log10 *p*-value) and the y-axis showing the SuSiE posterior inclusion probability (PIP). Alleles highlighted in red correspond to the top-ranked signals with a PIP greater that 0.8, with the five most significant labeled by allele name. The dashed horizontal line denotes the PIP = 0.8 threshold, commonly used as strong evidence for inclusion in the credible set.

**Supplementary Figure 2:**
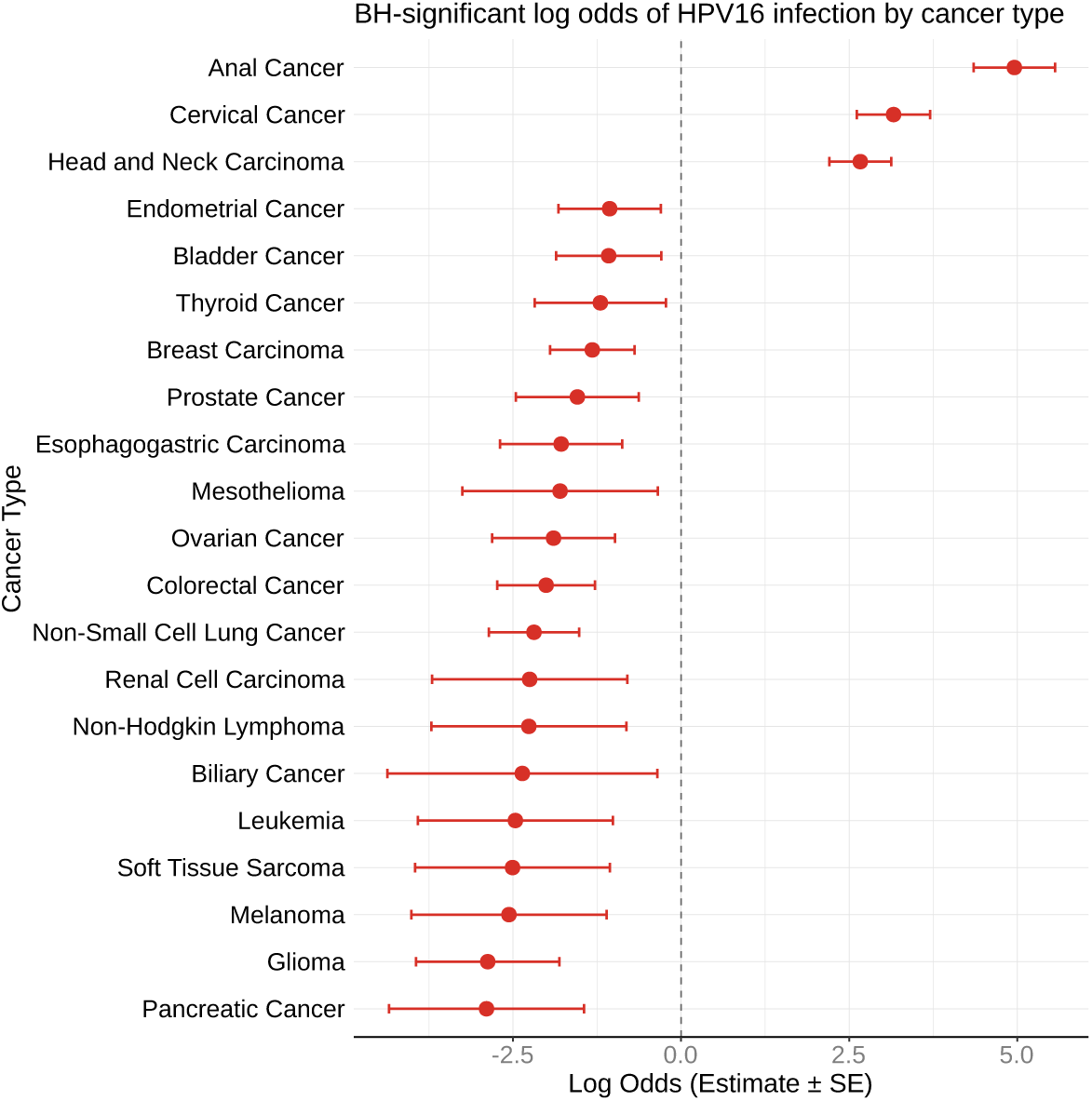
Associations of HPV16 Infection Across Cancer Types.

**Supplementary Table 1:**
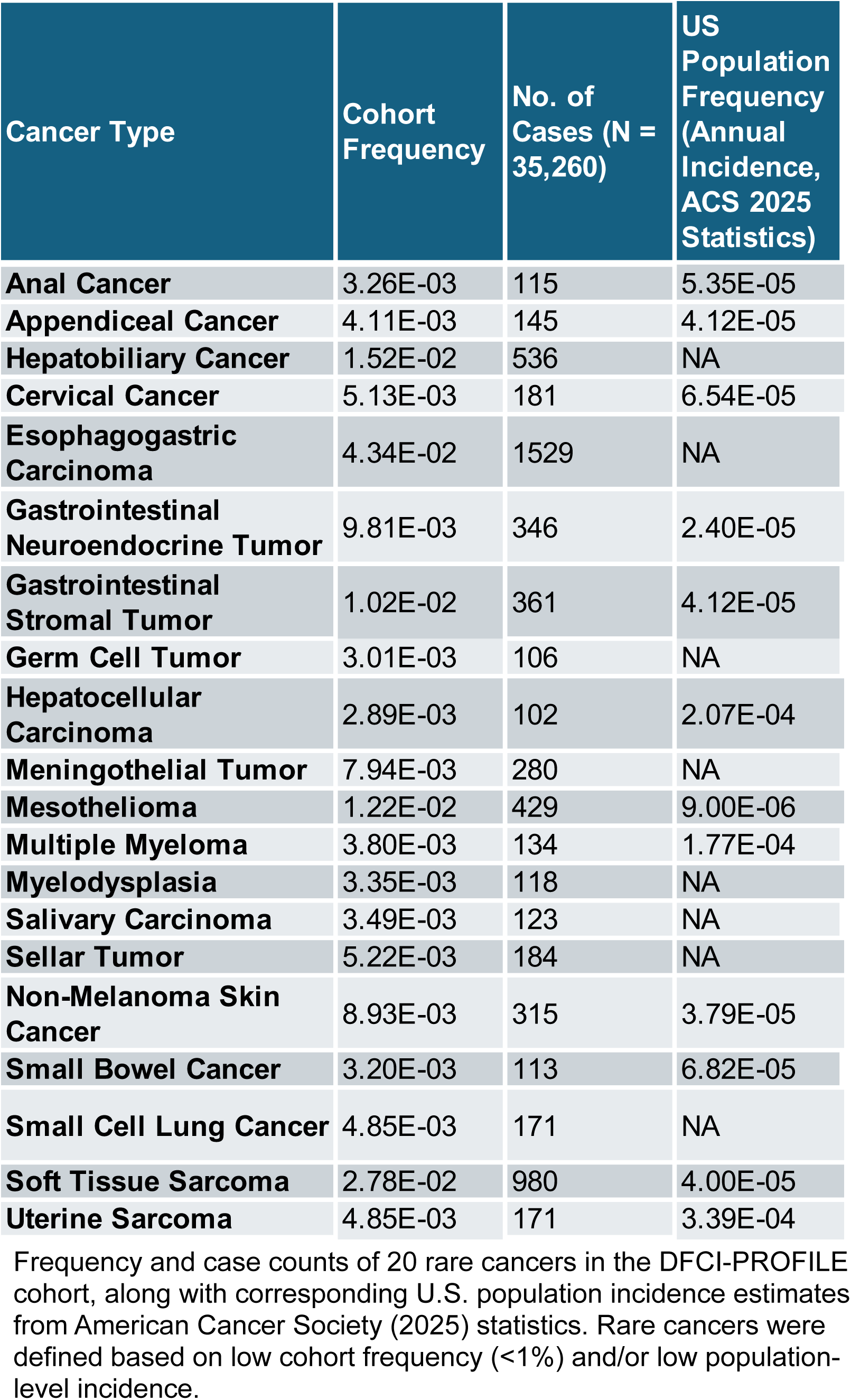
Frequency and case counts of rare cancers.

| Cancer Type | Cohort Frequency | No. of Cases (N = 35,260) | US Population Frequency (Annual Incidence, ACS 2025 Statistics) |
| --- | --- | --- | --- |
| **Anal Cancer** | 3.26E-03 | 115 | 5.35E-05 |
| **Appendiceal Cancer** | 4.11E-03 | 145 | 4.12E-05 |
| **Hepatobiliary Cancer** | 1.52E-02 | 536 | NA |
| **Cervical Cancer** | 5.13E-03 | 181 | 6.54E-05 |
| **Esophagogastric Carcinoma** | 4.34E-02 | 1529 | NA |
| **Gastrointestinal Neuroendocrine Tumor** | 9.81E-03 | 346 | 2.40E-05 |
| **Gastrointestinal Stromal Tumor** | 1.02E-02 | 361 | 4.12E-05 |
| **Germ Cell Tumor** | 3.01E-03 | 106 | NA |
| **Hepatocellular Carcinoma** | 2.89E-03 | 102 | 2.07E-04 |
| **Meningothelial Tumor** | 7.94E-03 | 280 | NA |
| **Mesothelioma** | 1.22E-02 | 429 | 9.00E-06 |
| **Multiple Myeloma** | 3.80E-03 | 134 | 1.77E-04 |
| **Myelodysplasia** | 3.35E-03 | 118 | NA |
| **Salivary Carcinoma** | 3.49E-03 | 123 | NA |
| **Sellar Tumor** | 5.22E-03 | 184 | NA |
| **Non-Melanoma Skin Cancer** | 8.93E-03 | 315 | 3.79E-05 |
| **Small Bowel Cancer** | 3.20E-03 | 113 | 6.82E-05 |
| **Small Cell Lung Cancer** | 4.85E-03 | 171 | NA |
| **Soft Tissue Sarcoma** | 2.78E-02 | 980 | 4.00E-05 |
| **Uterine Sarcoma** | 4.85E-03 | 171 | 3.39E-04 |

Frequency and case counts of 20 rare cancers in the DFCI-PROFILE cohort, along with corresponding U.S. population incidence estimates from American Cancer Society (2025) statistics. Rare cancers were defined based on low cohort frequency (<1%) and/or low population-level incidence.

**Supplementary Table 2:** Overview of study cohorts.

|  | DFCI-PROFILE | MSK-IMPACT | FinnGenn |
| --- | --- | --- | --- |
| N | 35,260 | 68,249 | 379,154 |
| Sex Prop. | 44% M / 56% F | 46% M / 54% F | 43% M / 57% F |
| Ancestry | Mixed Ancestry<br>EUR: 30,180<br>AMR: 1,946<br>AFR: 1,143<br>EAS: 878<br>SAS: 485<br>Other: 628 | Mixed Ancestry<br>EUR: 55,607<br>AMR: 832<br>AFR: 5,193<br>EAS: 3,651<br>SAS: 2,042<br>Other: 940 | Finnish (European) |
| Datatype | Oncopanel (germline imputed from tumor samples) | MSK-IMPACT panel (germline imputed from tumor samples) | Genotype chip |

Cohort characteristics for the GWAS meta-analysis.

**Supplementary Table 3:**
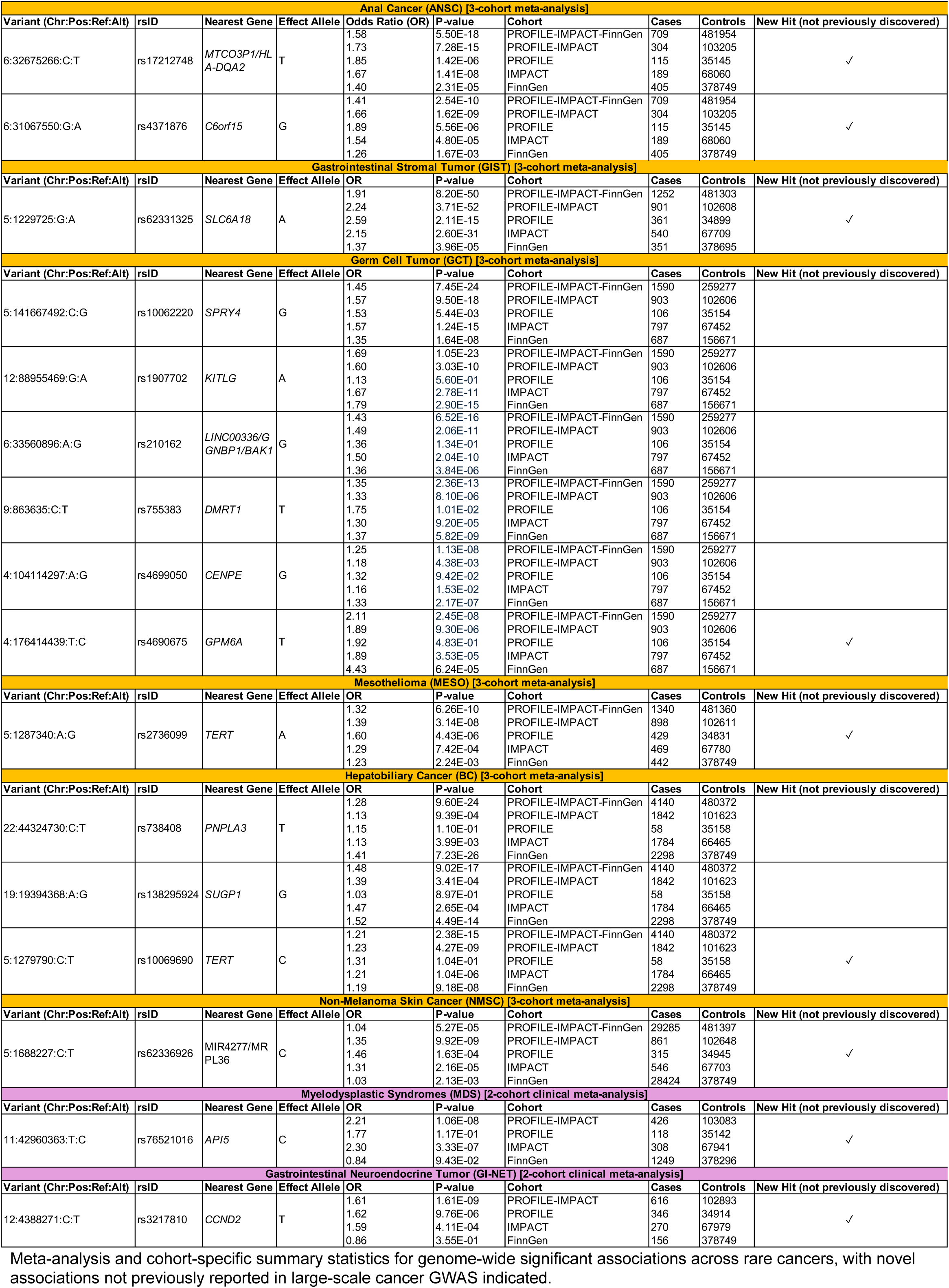
Genome-wide significant associations across rare cancers.

**Supplementary Table 4:**
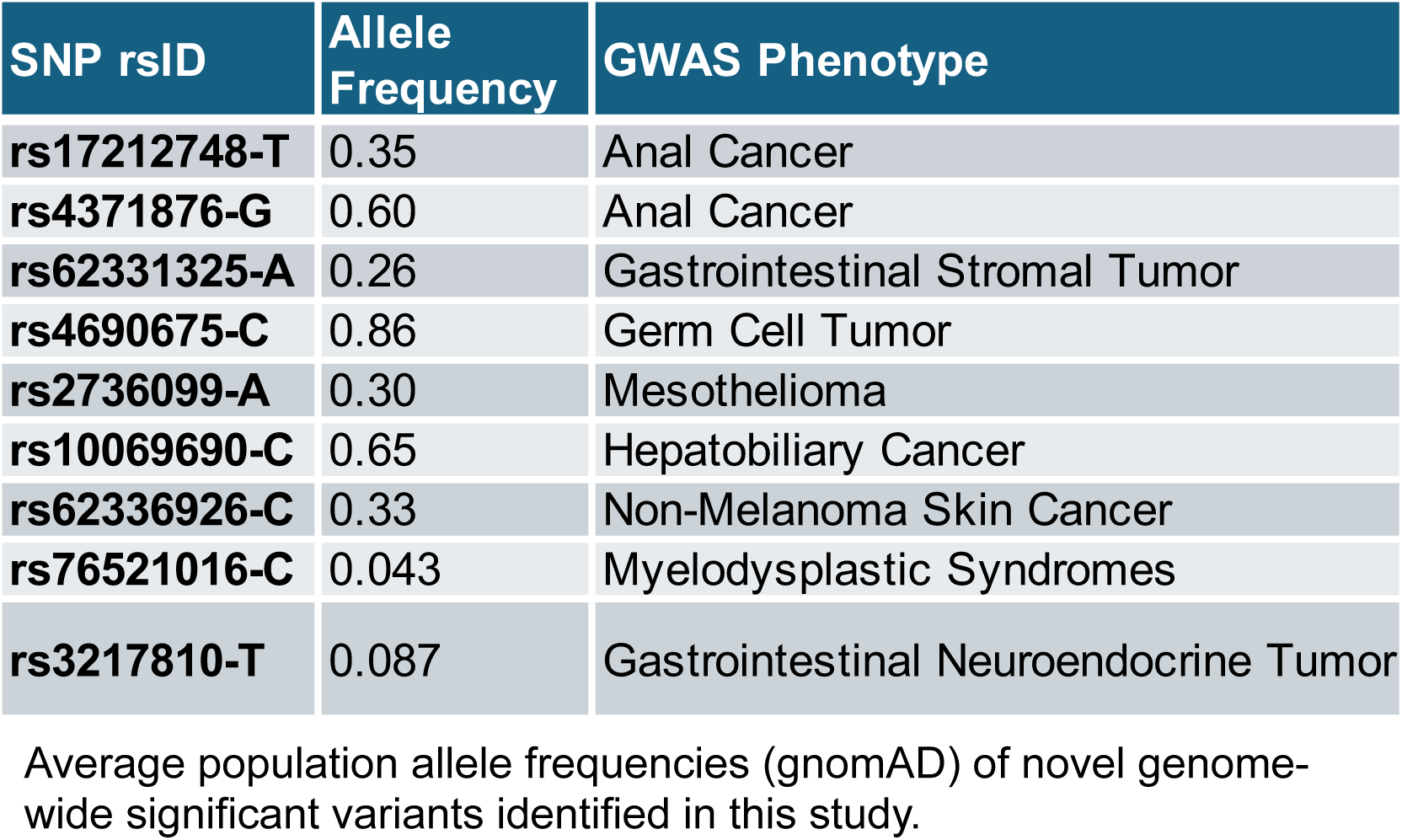
Allele frequencies of novel variants.

| SNP rsID | Allele Frequency | GWAS Phenotype |
| --- | --- | --- |
| rs17212748-T | 0.35 | Anal Cancer |
| rs4371876-G | 0.60 | Anal Cancer |
| rs62331325-A | 0.26 | Gastrointestinal Stromal Tumor |
| rs4690675-C | 0.86 | Germ Cell Tumor |
| rs2736099-A | 0.30 | Mesothelioma |
| rs10069690-C | 0.65 | Hepatobiliary Cancer |
| rs62336926-C | 0.33 | Non-Melanoma Skin Cancer |
| rs76521016-C | 0.043 | Myelodysplastic Syndromes |
| rs3217810-T | 0.087 | Gastrointestinal Neuroendocrine Tumor |

Average population allele frequencies (gnomAD) of novel genome-wide significant variants identified in this study.

**Supplementary Table 5:** Associations between germline variants and somatic mutations.

| Somatic Mutation | SNP | OR | P-value | Cancer Type | Cohort |
| --- | --- | --- | --- | --- | --- |
| KIT (SNV) | rs62331325-A | 2.21 | 6.50E-04 | GIST | PROFILE-IMPACT |
| KIT (SNV) | rs62331325-A | 2.67 | 4.90E-03 | GIST | PROFILE |
| KIT (SNV) | rs62331325-A | 1.92 | 3.60E-02 | GIST | IMPACT |
| PDGFRA (SNV) | rs62331325-A | 0.45 | 1.70E-02 | GIST | PROFILE-IMPACT |
| PDGFRA (SNV) | rs62331325-A | 0.31 | 5.00E-02 | GIST | PROFILE |
| PDGFRA (SNV) | rs62331325-A | 0.53 | 1.20E-01 | GIST | IMPACT |
| Quad Negative | rs62331325-A | 0.50 | 7.00E-02 | GIST | PROFILE |

Associations between somatic mutation status and lead germline variants across rare cancers, including results from PROFILE, IMPACT, and a combined PROFILE–IMPACT meta-analysis. Analyses were restricted to somatic mutations with frequency >5% in PROFILE across all ancestries.

**Supplementary Table 6:** Conditional analysis of GIST-associated variant and somatic driver mutations.

| Chr:Pos | SNP | Association (β) for SNP | P-value SNP | Association (β) for KIT | P-value KIT | Association (β) for PDGFRA | P-value PDGFRA | Association (β) for Quad Neg | P-value Quad Neg | Regression Model |
| --- | --- | --- | --- | --- | --- | --- | --- | --- | --- | --- |
| 5:1229725 | rs62331325-A | 0.65 | 0.24 | 8.91E-11 | 0.99 | NA | NA | NA | NA | PDGFRA ~ SNP + KIT \| GIST |
|  |  | 1.42 | 1.70E-02 | NA | NA | 2.03E-09 | 0.98 | NA | NA | KIT ~ SNP + PDGFRA \| GIST |
|  |  | NA | NA | 1.38 | 0.02 | 0.76 | 0.25 | NA | NA | SNP ~ PDGFRA + KIT \| GIST |
|  |  | NA | NA | 1.81 | 0.071 | 0.99 | 0.98 | 1.37 | 0.36 | SNP ~ PDGFRA + KIT + Quad Neg \| GIST |

Conditional analysis of the lead GIST-associated variant (rs62331325-A) and somatic driver mutations (KIT, PDGFRA) in the PROFILE cohort.

**Supplementary Table 7:** Association of GIST-associated variant with somatic mutations in non-GIST cancers.

| Chr:Pos | SNP | Association ($\beta$) for SNP | P-value SNP | Cancer Type | Somatic Mutation Tested |
| --- | --- | --- | --- | --- | --- |
| 5:1229725 | rs62331325-A | 0.92 | 0.87 | Melanoma | KIT |
|  |  | 0.82 | 0.71 | Glioma | PDGFRA |

Association between the lead GIST-associated variant (rs62331325-A) and somatic mutation status across non-GIST cancers in the PROFILE cohort (all ancestries).

**Supplementary Table 8:** Survival analysis of germline variant and KIT mutation status.

| Term | Cohort | Ancestry | Model | Coefficient ($\beta$) | Hazard Ratio | P-value |
| --- | --- | --- | --- | --- | --- | --- |
| SNP | PROFILE | ALL | Surv ~ SNP | 0.35 | 1.42 | 0.14 |
| SNP | IMPACT | ALL | Surv ~ SNP | 0.68 | 1.98 | 1.26E-03 |
| SNP | PROFILE | ALL | Surv ~ KIT + SNP | 0.35 | 1.42 | 0.14 |
| SNP | IMPACT | ALL | Surv ~ KIT + SNP | 0.06 | 1.85 | 4.66E-03 |
| SNP | PROFILE | ALL | Surv ~ KIT + SNP + KIT*SNP | -0.74 | 0.48 | 0.15 |
| SNP | IMPACT | ALL | Surv ~ KIT + SNP + KIT*SNP | 1.06 | 2.90 | 6.08E-02 |
| KIT*SNP | PROFILE | ALL | Surv ~ KIT + SNP + KIT*SNP | 1.40 | 4.06 | 1.53E-02 |
| KIT*SNP | IMPACT | ALL | Surv ~ KIT + SNP + KIT*SNP | -0.53 | 0.59 | 0.39 |

Cox proportional hazards models evaluating associations of rs62331325-A, KIT mutation status, and their interaction with overall survival in PROFILE and IMPACT cohorts.

**Supplementary Table 9:**
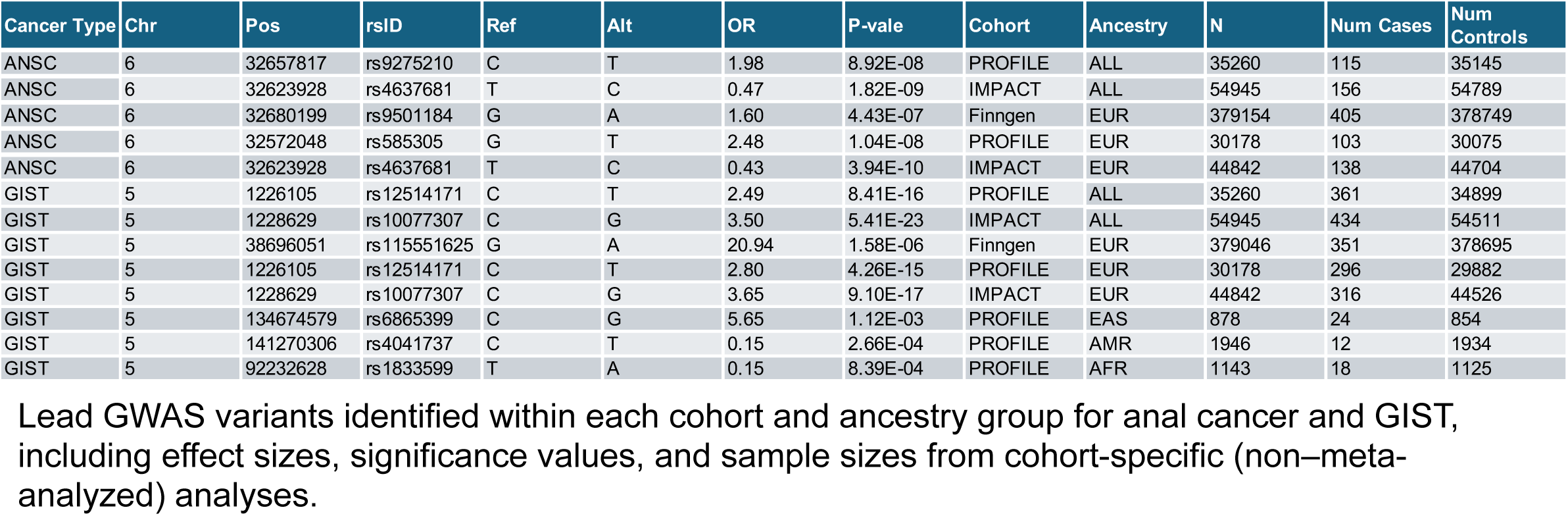
Cohort- and ancestry-specific lead GWAS variants.

| Cancer Type | Chr | Pos | rsID | Ref | Alt | OR | P-vale | Cohort | Ancestry | N | Num Cases | Num Controls |
| --- | --- | --- | --- | --- | --- | --- | --- | --- | --- | --- | --- | --- |
| ANSC | 6 | 32657817 | rs9275210 | C | T | 1.98 | 8.92E-08 | PROFILE | ALL | 35260 | 115 | 35145 |
| ANSC | 6 | 32623928 | rs4637681 | T | C | 0.47 | 1.82E-09 | IMPACT | ALL | 54945 | 156 | 54789 |
| ANSC | 6 | 32680199 | rs9501184 | G | A | 1.60 | 4.43E-07 | Finngen | EUR | 379154 | 405 | 378749 |
| ANSC | 6 | 32572048 | rs585305 | G | T | 2.48 | 1.04E-08 | PROFILE | EUR | 30178 | 103 | 30075 |
| ANSC | 6 | 32623928 | rs4637681 | T | C | 0.43 | 3.94E-10 | IMPACT | EUR | 44842 | 138 | 44704 |
| GIST | 5 | 1226105 | rs12514171 | C | T | 2.49 | 8.41E-16 | PROFILE | ALL | 35260 | 361 | 34899 |
| GIST | 5 | 1228629 | rs10077307 | C | G | 3.50 | 5.41E-23 | IMPACT | ALL | 54945 | 434 | 54511 |
| GIST | 5 | 38696051 | rs115551625 | G | A | 20.94 | 1.58E-06 | Finngen | EUR | 379046 | 351 | 378695 |
| GIST | 5 | 1226105 | rs12514171 | C | T | 2.80 | 4.26E-15 | PROFILE | EUR | 30178 | 296 | 29882 |
| GIST | 5 | 1228629 | rs10077307 | C | G | 3.65 | 9.10E-17 | IMPACT | EUR | 44842 | 316 | 44526 |
| GIST | 5 | 134674579 | rs6865399 | C | G | 5.65 | 1.12E-03 | PROFILE | EAS | 878 | 24 | 854 |
| GIST | 5 | 141270306 | rs4041737 | C | T | 0.15 | 2.66E-04 | PROFILE | AMR | 1946 | 12 | 1934 |
| GIST | 5 | 92232628 | rs1833599 | T | A | 0.15 | 8.39E-04 | PROFILE | AFR | 1143 | 18 | 1125 |

Lead GWAS variants identified within each cohort and ancestry group for anal cancer and GIST, including effect sizes, significance values, and sample sizes from cohort-specific (non–meta-analyzed) analyses.

**Supplementary Table 10:** Association of ANSC variants with HPV status.

| Chr:Pos | SNP | OR | P-value | Ancestry | Cohort |
| --- | --- | --- | --- | --- | --- |
| 6:32572048 | rs585305-T | 1.36 | 2.11E-04 | All | PROFILE only |
|  |  | 1.44 | 3.19E-05 | EUR |  |
| 6:32675266 | rs17212748-T | 1.19 | 2.40E-02 | All | PROFILE-IMPACT-FinnGen |
|  |  | 1.20 | 2.60E-02 | EUR |  |

Association of lead ANSC-associated variants from meta-analysis and cohort-specific analyses with HPV viral read status across all patients and cancer types in the PROFILE cohort, adjusted for cancer type.

**Supplementary Table 11:** Association of ANSC variant with HPV status across cancer types.

| Cancer Type | Chr:Pos | SNP | OR | P-value | N | N Cases |
| --- | --- | --- | --- | --- | --- | --- |
| Anal Cancer | 6:32572048 | rs585305-T | 1.68 | 3.80E-02 | 115 | 83 |
| Cervical Cancer |  |  | 2.00 | 2.20E-04 | 181 | 60 |
| Head and Neck Carcinoma |  |  | 1.15 | 1.30E-01 | 599 | 192 |
| Colorectal Cancer |  |  | 1.00 | 9.90E-01 | 3794 | 189 |

Association of the lead ANSC-associated variant (rs585305-T) with HPV viral read status across HPV-related cancer types in the PROFILE cohort (European ancestry), stratified by cancer type.

**Supplementary Table 12:** Associations between MDS variant and hematologic traits.

| Blood Test | Association<br>($\beta$) for Blood<br>Test | P-value |
| --- | --- | --- |
| Hemoglobin (HGB) | 9.53E-03 | 0.49 |
| Hematocrit (HCT) | 0.017 | 0.67 |
| Red Blood Cells<br>(RBC) | 2.76E-03 | 0.56 |
| Platelets (PLT) | 1.22 | 0.17 |
| White Blood Cells<br>(WBC) | 6.80E-02 | 5.30E-02 |
| Absolute<br>Neutrophils (Neuts) | 77.72 | 4.56E-03 |

Associations between the MDS risk variant rs76521016-C and routine hematologic measurements.

**Supplementary Table 13:**
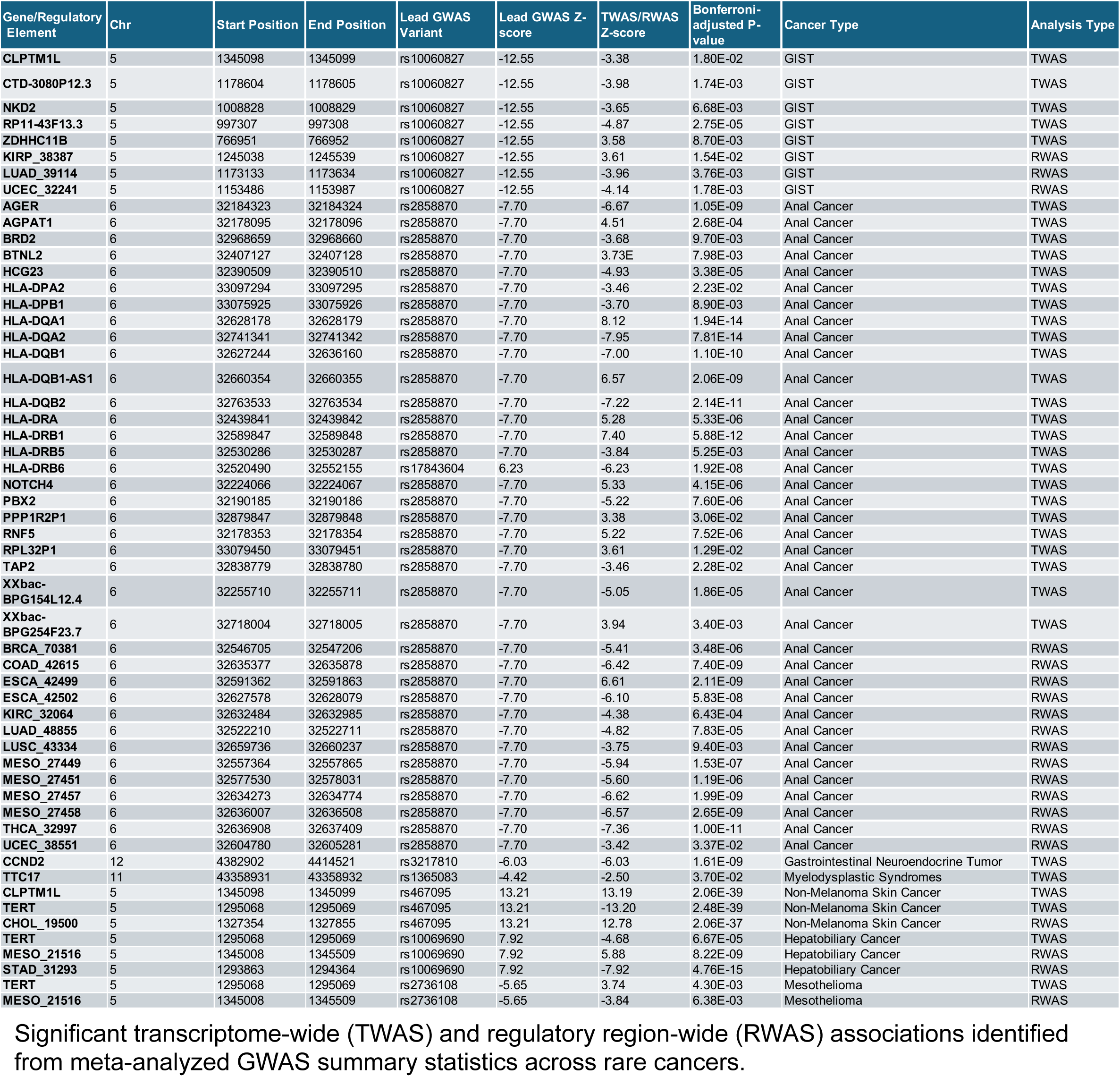
Significant TWAS and RWAS associations.

**Supplementary Table 14:** Frequency of HPV strains in the PROFILE cohort.

| HPV Strain Type | Cohort Frequency | No. of Samples |
| --- | --- | --- |
| Human papillomavirus 16 | 3.27E-01 | 467 |
| Human papillomavirus 18 | 1.75E-02 | 25 |
| Human papillomavirus 10 | 4.90E-03 | 7 |
| Human papillomavirus 4 | 4.20E-03 | 6 |
| Human papillomavirus type 6b | 4.20E-03 | 6 |
| Human papillomavirus 9 | 1.40E-03 | 2 |
| Human papillomavirus type 34 | 1.40E-03 | 2 |
| Human papillomavirus type 49 | 1.40E-03 | 2 |
| Human papillomavirus type 53 | 1.40E-03 | 2 |
| Human papillomavirus | 7.00E-04 | 1 |
| Human papillomavirus 112 | 7.00E-04 | 1 |
| Human papillomavirus 116 | 7.00E-04 | 1 |
| Human papillomavirus 129 | 7.00E-04 | 1 |
| Human papillomavirus 175 | 7.00E-04 | 1 |
| Human papillomavirus 32 | 7.00E-04 | 1 |
| Human papillomavirus type 26 | 7.00E-04 | 1 |
| Human papillomavirus type 54 | 7.00E-04 | 1 |

Frequency and sample counts of detected HPV strains in the DFCI-PROFILE cohort based on viral read analysis of tumor sequencing data.

